# Cognitive Flexibility and Decision-Making in Anxiety and Depression: Meta-Analytic Evidence Facilitated by Machine-Learning Screening

**DOI:** 10.64898/2026.05.14.26353209

**Authors:** Juan Balcazar, Brian Albanese, Taylor Rymer, Madelynn Davis, Sofia Campos, Madeline Polimerou, Emma Abel, Jessie Shapley, Ilanit Algranatti, Hope Wood, Hadley Smith, Kate Hankamer, Joseph M. Orr

## Abstract

The ability to adjust to changing environments (cognitive flexibility) and optimal decision-making are pivotal brain functions that govern successful human behavior. Anxiety and depressive disorders are strongly pervasive psychiatric conditions across the lifespan that profoundly disrupt mechanisms of attention, working memory, and decision-making. Although existing task evidence documents impaired decision-making and flexibility outcomes for both anxiety and depression, there is a growing need to systematically evaluate the role of anxiety and depression and to quantitatively compare the effects of these disorders on these domains. In the present study, we conducted a meta-analysis of anxiety and depression on decision-making and cognitive flexibility. We utilized a random-effects approach, given that a large amount of between-subject heterogeneity was anticipated. Given the scope of this meta-analysis, we used the machine learning tool ‘ASReview’ to more efficiently conduct meta-analytic screening. Across all outcomes, results showed anxiety and depression were associated with reduced cognitive flexibility and decision-making. These effect sizes were then tested for significance using a fixed-effects (plural) model. Subgroup analyses revealed no significant differences between anxiety and depression for either decision-making or flexibility outcomes, consistent with a transdiagnostic perspective. Results are contextualized in light of the biopsychosocial model and potential transdiagnostic factors.

## MAIN

Anxiety and depressive disorders are pervasive across the lifespan (Bandelow & Michaelis, 2015; Hasin et al., 2018; Merikangas et al., 2010), with a lifetime prevalence in the U.S. of 20% and 30%, respectively (Hasin et al., 2018; LeDuke et al., 2023). Around the globe, anxiety has a prevalence of 4% (Institute for Health Metrics and Evaluation [IHME], 2020), and depression has a prevalence of 3.8% (Institute for Health Metrics and Evaluation [IHME], 2020). Anxiety disorders reflect heightened distress, arousal, and vigilance in the face of some potential threat (Hamm, 2020; Shackman & Fox, 2021) and are often linked with disproportionate responses to life stressors, restlessness, irritability, and persistent worry (Kenwood et al., 2022; LeDuke et al., 2023; Maina & Rossi, 2016). Depressive disorders—which include major depressive disorder (MDD), disruptive mood dysregulation, premenstrual dysphoric disorder, and persistent depressive disorder—are characterized by sad and irritable moods, leading to heightened stress and cognitive impairment (Maina & Rossi, 2016). MDD is also characterized by diminished interest and pleasure (anhedonia), low energy, or inefficiency when engaging in daily tasks (American Psychological Association [APA], 2013; LeDuke et al., 2023). Considering the widespread prevalence and significant impairment associated with these disorders, it is imperative to understand the consequences of these conditions in relation to human cognition.

The relationship between anxiety and depression is notably complicated. Individuals frequently show co-occurring symptoms (i.e. *comorbidity*; Jacobson & Newman, 2014; LeDuke et al., 2023; Mineka et al., 1998). Interestingly, anxiety predicts later depression more than depression predicts later anxiety (Doering et al., 2019; Jacobson & Newman, 2014; Merikangas et al., 2003). Reflecting this complex relationship, anxiety and depression exhibit *shared* and *distinct* traits (Bishop & Gagne, 2018; LeDuke et al., 2023). The pattern of high comorbidity alongside separable features motivated efforts to model their relationship. An early endeavor, the tripartite theory proposed that negative affect represents the shared trait between anxiety and depression, explaining their comorbidity (Clark & Watson, 1991; for a review see Anderson & Hope, 2008). As new evidence emerged, Clark and Watson (2006) theorized that anxiety and depression comprise distinct underlying fear and distress-based subfactors yet map to a shared latent factor. This structure was later incorporated into the Hierarchical Taxonomy of Psychopathology (HiTOP) model (Kotov et al., 2017). Specifically, the model posits that anxiety and depression disorders share a latent *internalizing* factor; importantly, major depressive disorder and generalized anxiety disorder load onto a *distress* subfactor characterized by negative affect, whereas other anxiety disorders, including panic disorder, load onto a *fear* subfactor characterized by autonomic arousal and broad physiological fear responses. Collectively, this evidence suggests anxiety and depression share common latent factors, yet it remains unclear whether they exert shared or perhaps distinct effects on cognitive functioning.

Empirical investigations have shown that anxious and depressed moods are linked to disruptions in basic cognitive mechanisms, including attention (Berggren & Derakshan, 2013; Ortega et al., 2017; Shi et al., 2019; Thomas et al., 1999; Du et al., 2022), working memory (Gazzaley, 2011; Moran, 2016), and affective control (Deveney & Deldin, 2006; Liu & Wang, 2014). Building upon these foundational deficits, research has also shown that anxiety and depression impair higher-order cognitive processes, specifically cognitive flexibility (Deveney & Deldin, 2006; Murphy et al., 2012; Park & Moghaddam, 2017) and decision-making (Gao et al., 2021; Hartley & Phelps, 2012; Mueller et al., 2010; Wu et al., 2013). Unlike attention and working memory, which serve as cognitive building blocks, flexibility and decision-making represent integrative processes that directly translate into adaptive behavior. We focus on these domains because they are particularly consequential for real-world functioning. For instance, deciding when to purchase stocks requires optimal decision-making, whereas adapting an investment strategy in response to market fluctuations requires cognitive flexibility. Understanding how anxiety and depression impact these higher-order processes is therefore critical for characterizing functional impairment in these disorders.

Despite extensive research, important gaps remain in understanding how anxiety and depression affect cognitive flexibility and decision-making outcomes. First, prior meta-analytic work has examined specific risk factors of anxiety and depression (e.g. Buur et al., 2025, Jacobson & Newman, 2017, Hong & Cheung, 2015), yet these meta-analytic efforts rely on self-report (e.g. Buur et al., 2025; Hong & Cheung, 2015). Although self-report and task-based cognitive measures capture related constructs, they are empirically distinct (Duckworth & Kern, 2011; Toplak et al., 2013), with self-report susceptible to social desirability bias and less able to capture autonomic cognitive responses (Dang et al., 2020; Hohl & Dolcos, 2024; Howlett et al., 2021). Thus, prior meta-analytic reliance on self-report leaves the task-based cognitive profiles of anxiety and depression underexamined. Existing meta-analytic evidence from single-task behavioral data sets reports inconclusive effects and calls for additional synthesis (Bishop & Gagne, 2018; Huys et al., 2013). Second, considerable heterogeneity in task designs complicates interpretation. For instance, the decision-making construct has been assessed using diverse methodologies (e.g., Iowa Gambling Task, risk-taking tasks, effort expenditure tasks, social decision-making paradigms); similarly, cognitive flexibility has been measured through multiple paradigms, including card-sorting tasks, reversal learning, and task-switching paradigms. It therefore remains unclear whether anxiety and depression consistently impair performance across varied task-based measures, a question a comprehensive meta-analysis is able to address. Third, sub-domains within decision-making remain under-examined. Notably, while social decision-making is clinically relevant to both disorders, its relationship to anxiety and especially depression is poorly understood compared to traditional decision-making tasks (Safra et al., 2019; Destoop et al. 2012), making it a priority for meta-analytic investigation.

In summary, anxiety and depression are jointly studied because of their high prevalence, public health impacts, and comorbidity. Their complex relationship is underscored by models that show a shared *internalizing* latent factor and separate subfactors (e.g. HiTOP model). Taken together, a meta-analysis is warranted to synthesize the growing literature and clarify disorder-specific effects on cognitive flexibility and decision-making across heterogeneous task-based assessments. We next review task evidence for anxiety and depression on decision-making and cognitive flexibility outcomes.

### Task evidence for Decision-making outcomes

Because decisions can have adverse consequences, the brain must weigh potential rewards against threats (Broman-Fulks et al., 2014; Kahneman, 2003). Anxiety in particular has been linked to disruptions in this reward-threat weighting process. For instance, individuals with high trait anxiety show impaired decision-making under risk, avoiding options linked to recent losses (Mueller et al., 2010) and becoming risk-averse after gains (Giorgetta et al., 2012).

Although depression has also been associated with reduced risk-taking—particularly in transient induced sad states(Yuen & Lee, 2003)—evidence at the disorder level remains more mixed (Huys et al., 2013), whereas anxiety shows more consistent risk avoidance (Ortega et al., 2017). Anxiety-related risk avoidance also appears in social contexts, reflecting decision-making patterns characterized by information restriction and avoidance of uncertain social outcomes.For instance, individuals with social anxiety disorder often exhibit social risk aversion by minimizing the immediate probability of rejection; specifically, they are less willing to express romantic interest if a face-to-face meeting is expected soon, seek less information to avoid potentially threatening feedback, and show greater apprehension about future social events (Rozen & Aderka, 2022). Anxiety is further linked to altered learning rates, whereas evidence for such alterations in major depressive disorder is mixed (Huys et al., 2013; Pulcu & Browning, 2019). Specifically, anxious individuals fail to adjust learning rates optimally, particularly after negative outcomes (Browning et al., 2015), are insensitive to loss frequency, and respond poorly to infrequent losses (Mueller et al., 2010). Together, increased risk avoidance and altered learning rates in anxiety can disrupt decision-making by limiting the information that is sampled (see Bishop & Gagne, 2018). Therefore, we hypothesize that anxiety is associated with moderate to large, negative effects on decision-making and, relative to depression, yields larger effect sizes.

***H1a: The effects of anxiety disorders on decision-making tasks are moderate to large and negative*.**

***H1b: The effects of anxiety disorders on decision-making tasks are larger as compared to depression*.**

### Task evidence for Flexibility outcomes

Under rapidly changing conditions, the brain must balance flexible control (e.g., seizing new rewards) with stable control (e.g., resisting distraction) (Eppinger et al., 2021; Goschke & Dreisbach, 2008). Disruption of this balance can produce perseveration, where goals are rigidly maintained, or excessive updating, and where behavior becomes overly driven by distraction (Del Giudice & Crespi, 2018). Attentional control is central to cognitive flexibility. Attentional Control Theory proposes that anxiety impairs cognition by reducing the ability to inhibit distractors, shift attention, and update working memory (Eysenck et al., 2007; Ortega et al., 2017), and that these mechanisms strongly shape flexibility.

Supporting work shows that anxiety is linked to deficits in attention processing, especially inhibition and switching (Du et al., 2022; Lee & Orsillo, 2014; Shi et al., 2019). However, Barthel and colleagues (2022) note that flexibility impairments in anxiety are task-specific; whereas, anxiety effects are most consistent for the Wisconsin Card Sorting Test, findings for the Trail Making Test are mixed (Barthel et al., 2022; Baussay et al., 2024; Waldstein et al., 1997). Other work indicates that effects depend on anxiety *severity*. Moderate anxiety may narrow attention in ways that support performance; for example, high trait anxiety in non-clinical samples has been associated with better attention and working-memory performance (Dotson, 2014). The suggested explanation is that whereas anxiety may reduce processing efficiency, it may increase compensatory effort and thus preserve overall performance (Eysenck & Calvo, 1992), with impact varying by task demands such as high-versus low-fatigue conditions (O’Shea et al., 2016). In panic disorder, evidence for flexibility deficits is also mixed (Giomi et al., 2021). At the cognitive level, when switching between tasks, individuals with anxiety demonstrate elevated switch costs (delayed reaction times when transitioning to a new task), particularly when disengaging from attentionally demanding tasks to easier ones (Gustavson et al., 2017; Park & Moghaddam, 2017; Remijnse et al., 2013). This pattern suggests that anxiety-related flexibility deficits reflect difficulty disengaging from effortful tasks rather than general cognitive impairment.

Like anxiety, depression is also associated with impaired cognitive flexibility, but the behavioural patterns that individuals show differ. Depressed individuals show elevated switch costs (Park & Moghaddam, 2017; Remijnse et al., 2013) and, critically, higher overall error rates on flexibility tasks compared to controls (Zheng et al., 2024), suggesting broader difficulties with switch efficiency rather than task-specific disengagement problems. Taken together, although both disorders impair flexibility, they appear to do so through distinct mechanisms. That is, anxiety-related deficits are linked to broader attentional disruption, affecting threat detection and attentional orientation independent of emotional stimuli (Eysenck et al., 2007; Ortega et al., 2017), and manifest primarily as difficulty disengaging from demanding tasks. In contrast, depression-related deficits are mediated by emotional valence (Deveney & Deldin, 2006; Mogg & Bradley, 2005; Whitmer & Gotlib, 2012, 2013) and appear as general switch inefficiency with elevated error rates. Depression may thus show stronger and more consistent impairments in flexibility, driven by emotion-related attention deficits, negative affect, and ruminative thought patterns, all of which have been associated with reduced flexibility in prior work (Hsieh & Lin, 2019). Given the mixed findings for anxiety(i.e., varied effects by task type, severity, and absent or even beneficial at moderate levels) and the more consistent pattern of impairment in depression, we hypothesize that depression shows larger and more reliable effects on cognitive flexibility than anxiety. Overall, anxiety does affect cognitive flexibility, but its effects are task-and context-dependent, and can be beneficial at moderate levels.

***H2a: The effects of depressive disorders on cognitive flexibility tasks are moderate to large and negative*.**

***H2b: The effects of depressive disorders on cognitive flexibility tasks are larger compared to anxiety disorders*.**

## Methods

### Meta-analytic criteria

Studies were included in the meta-analysis if they met four criteria: a-c. See Table 1.

**Table 1:**
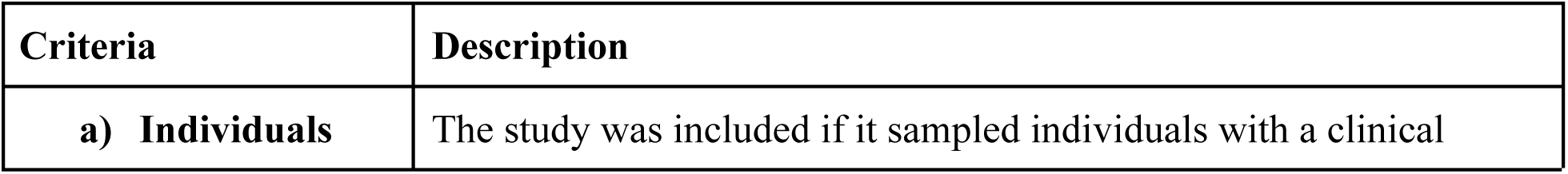
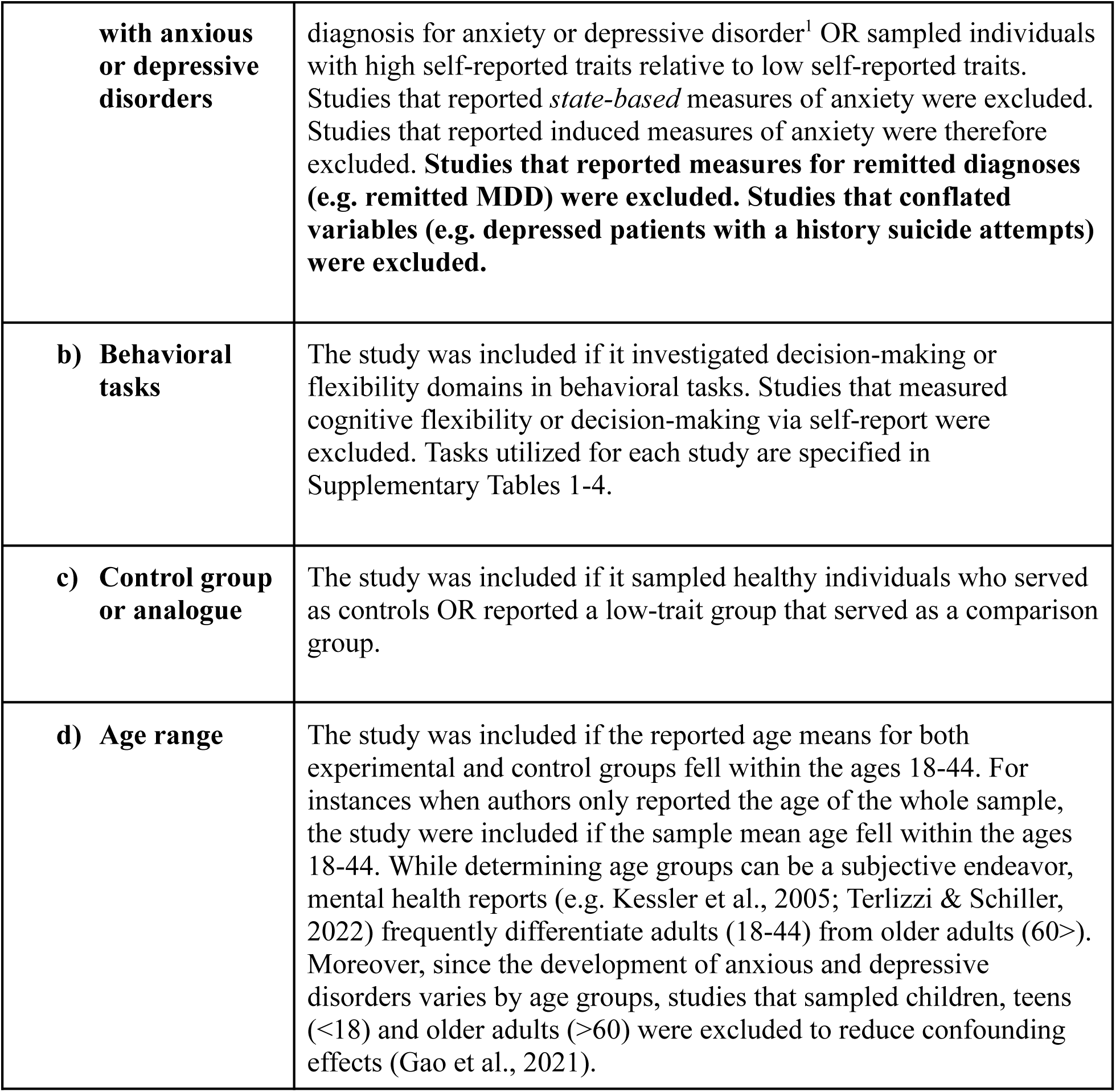
Meta-analytic Criteria for the present study.

### Literature search

A systematic review of the literature was conducted to identify articles for inclusion. Consistent with established practices in systematic review for mental health topics, multiple databases were searched (Higgins et al. 2019). The following databases were searched: *PUBMED*, *APA PsycInfo*, *ERIC*, and *Academic Search Ultimate*. The following search terms were used: “anxiety AND decision-making”, “anxiety AND cognitive flexibility”, “depression AND decision-making”, “depression AND cognitive flexibility”. The APAPsycInfo, ERIC, and Academic Search Ultimate databases (henceforth referred to *APAPsyc)* were simultaneously queried and data retrieved as one file. PUBMED was queried and retrieved as one file. To allow for an exhaustive search, queries were not bounded by publication date and no custom range was utilized^2^.

### Screening strategy

The search yielded approximately 23,587 records. First, duplicate records were removed using an automated tool (*N*= 3391). Initially, trained experimenters inspected the abstract, title, or main text (i.e. screening) of 4077 articles for the meta-analytic criteria shown in Table 1. Because of the scale of the present study, AI-assisted screening was introduced mid-project. We used the machine-learning tool *ASReview Lab* (version 1.6.3; van de Schoot et al., 2021), which uses active learning to learn, in real-time, user inputs and then displays the most relevant title and abstract, rank-ordered by expected relevancy; the user marks whether an abstract is relevant or not. In this way, ASReviewLab rank-ordered the remaining unscreened records, prioritizing those most likely to be relevant. Low-relevance items remained at the bottom of the AI ranking and were not manually screened. Because the machine-learning tool did not exclude or remove any records, these unscreened items remained part of the overall set of records submitted to the screening process. As a result, the total number of screened records equaled the total number of identified records (see Figure 2: PRISMA). Overall, the present study represents one of the more extensive systematic screenings in clinical/cognitive psychology literature, in part due to the combination of human review and machine-assisted screening. See Figure 1 for the setup, operation, and verification workflows involving the machine-learning tool.

**Figure 1:**
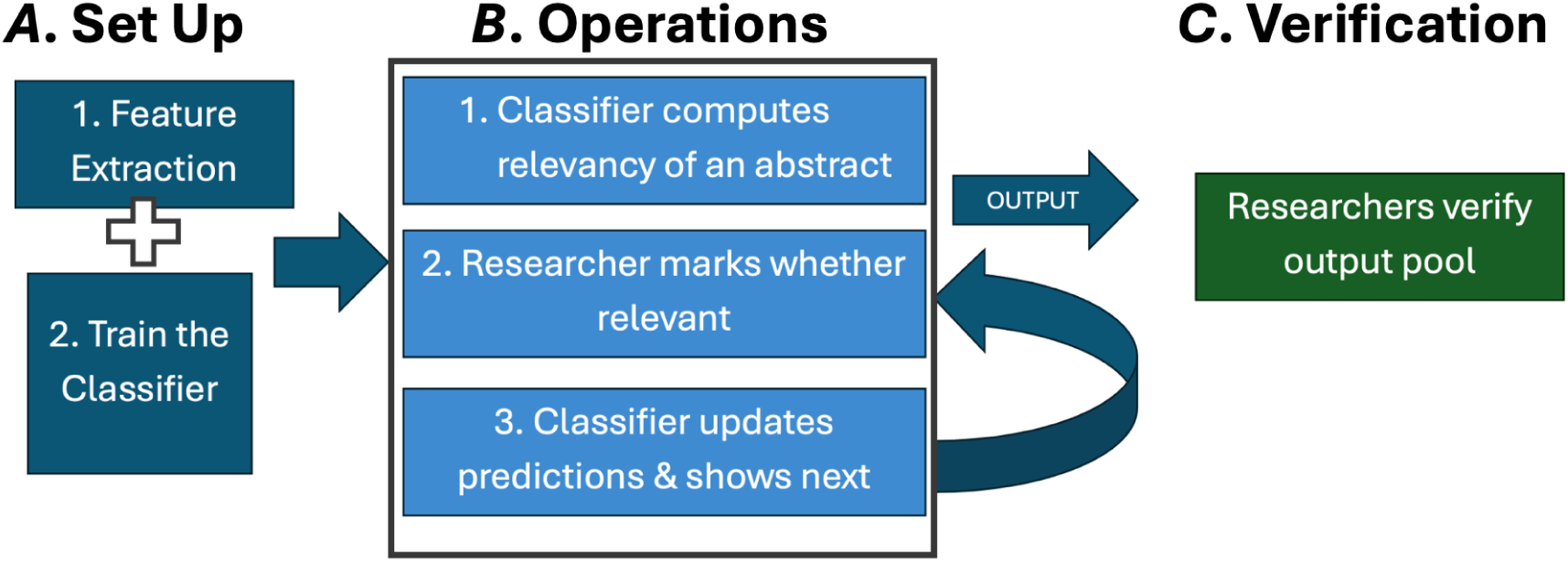
A. Set-up, Stage at which model parameters were selected which are also the defaults for ASReview lab,. Model: Naive Bayes; Query strategy: Maximum; Balance strategy: Dynamic resampling (double); Feature extraction: TF-IDF. The classifier was trained on 4 relevant articles and 5 irrelevant articles to train the model (i.e. precursor knowledge^3^). B. Operations, The classifier returns a list of abstract ordered from most relevant to least relevant. Researchers mark whether it is relevant or irrelevant based on the meta-analytic criteria. The classifier updates its predictions and serves the next relevant abstract. C. Verification: All abstracts go through detailed manual review to determine their adequacy or statistical viability for meta-analytic inclusion.

**Figure 2.**
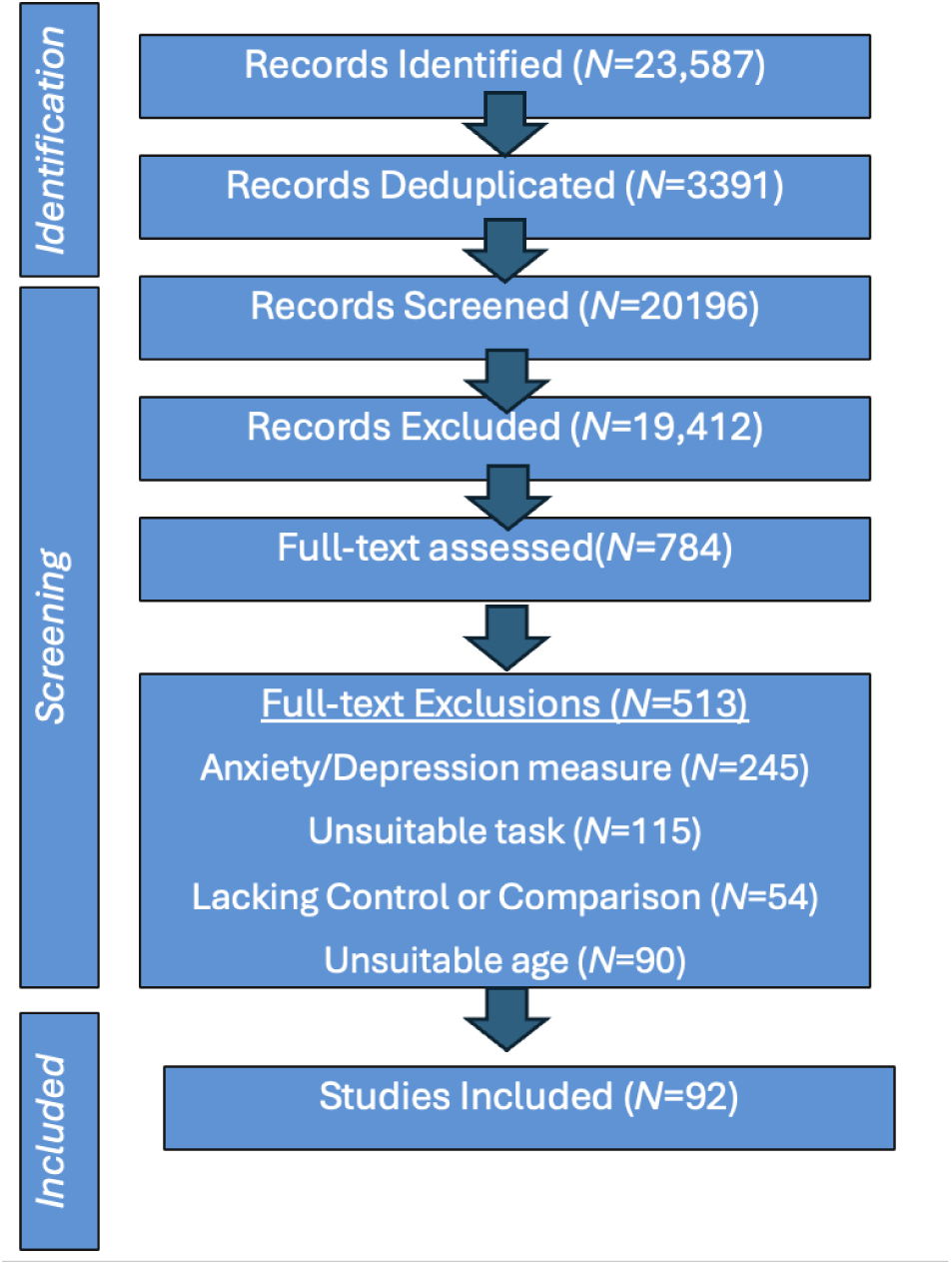
PRISMA flow diagram for study selection. A total of 23,587 records were identified through database searches, and deduplicated (N=3391). These remaining records were screened at the title/abstract level using a combination of manual and AI-assisted screening. A total of 784 full-text articles were assessed for eligibility across four outcome domains (Anxiety–Decision-making, Anxiety–Flexibility, Depression–Decision-making, and Depression–Decision-Making). Following full-text assessment, exclusions were categorized as follows: 254 for unsuitable anxiety/depression measures, 115 for unsuitable tasks, 54 for unsuitable or missing healthy control groups or a low-trait group, and 90 for age-related ineligibility. Two articles were paywalled. The final number of studies included in the meta-analysis was 92 unique studies (contributing to 100 effect sizes).

#### Stopping rule

In the present study, a stopping rule was determined apriori before conducting AI-assisted screening with *ASReviewLab*. Given that statistically derived stopping rules (e.g. estimate derivations of relevant papers in a training set) can often be impractical (Campos et al., 2024), stopping rules in the form of heuristics *(e.g. time-based stopping rule)* have been favored by prior researchers. Effective time-based stopping rules should address the limited screening time available to researchers while maximizing the number of relevant records that could be found. In the present study, because trained researchers working in shifts completed 20 hours of AI-assisted time in approximately 1.5 to 2 weeks, 20 hours served as a good balance between the cost of screening and the cost of overlooking relevant papers. (see, Cohen, 2011). To benefit from data-driven approaches, the present study utilized a *mixed* strategy that employs a time-based stopping rule alongside a data-driven heuristic (i.e. consecutive irrelevant records) as a stopping rule. For the data-driven stopping rule, the 50 consecutive irrelevant papers were chosen as recommended by Boetje and van de Schoot (2024). Thus, for each retrieved database (e.g. a PUBMED database for anxiety and cognitive flexibility), screening was considered complete when the predetermined number of 20 hours for total AI-assisted screening time was met (time-based stopping rule), unless the predetermined threshold of 50 consecutive irrelevant papers was met before that time (data-driven rule). For further methodological discussion of mixed and data-driven heuristic approaches, the reader is referred to Campos et al., 2024; Hamel et al., 2021. Supplementary Table 5 lists stopping outcomes for each database.

We also followed recommendations to visually inspect the model recall plots for each screening instance. For all instances, we saw a discernible plateau in the recall trajectory, indicating that the probability of identifying additional relevant records becomes small (Boetje & van de Schoot, 2024; Ferdinands et al., 2023). This provided additional confidence that screening stopped appropriately, reflecting the low likelihood of identifying further relevant abstracts.

### Machine Learning Model: ASReview lab

In the present study, the primary benefit of utilizing AI-assisted screening is to decrease the likelihood of overlooking a relevant record without burdening computational cost. Thus, we considered the efficiency of the feature extractor and model parameters for suitability in the present study. The model parameters selected, which are also the defaults for ASReview lab, were the following: Model: Naive Bayes; Query strategy: Maximum; Balance strategy: Dynamic resampling (double); Feature extraction: TF-IDF.

To ensure high-quality data input for the classifier, *ASReview* documentation and recommendations were followed. First, duplicates and incomplete cases (e.g. records without an abstract or title) were removed. In addition to reducing the screening burden (Hamel et al., 2021), deduplication ensures high-quality data input for the classifier. The remaining records were then imported into *ASReview* and the aforementioned model parameters selected. For all instances (anxiety and decision-making, anxiety and cognitive flexibility, depression and decision-making, depression and cognitive flexibility), we selected 4 relevant articles and 5 irrelevant articles to train the model (i.e. precursor knowledge^4^). The feature extractor and model training, ranking the records in the dataset, completed rapidly (within 30 seconds or less for all instances). Researchers underwent comprehensive training to understand the meta-analytic criteria and appropriately operate ASReview. Co-authors that underwent training led by author JB are as follows: M.D., S.C., I.A., M.P., T. R. Next, researchers inspected abstract text for the meta-analytic criteria (Table 1); an abstract was labeled *relevant* if the article satisfied both Criteria *A* and *B* as described in Table 1. Because abstracts often prioritize brevity, it is probable that additional criteria important to the present study (e.g. Criteria *C*, the use of healthy controls or control analogues) may not be reported in the abstract text. Thus, if an abstract text satisfied Criteria *A* and *B*, but did not address the remaining criteria, the abstract was marked *relevant* alongside a documented note indicating full-text inspection was warranted. Upon reaching the stopping rule (Supplementary Table 5), all abstracts labeled *relevant* were exported to a separate spreadsheet so that trained researchers could closely inspect the *article text* for remaining criteria (i.e. full-text assessment). Following this manual inspection of the article text, if a study meets all criteria (Table 1), researchers extracted relevant statistics for statistical conversion. Additional articles were identified by consulting the reference section of studies (N=3).

### Statistical conversion

For articles that met all meta-analytic criteria, we inspected the article’s results section (including supplementary data and supplementary files) in order to convert the reported statistics to Hedges’ *g*. Hedges’ *g* describes the standardized mean difference correcting for small sample bias, and allows for meaningful comparison across studies of different designs and methods. We used the R package ‘esc’(Lüdecke et al., 2019) to compute Hedges’ *g* and its standard error using the statistics reported in each study. The esc package affords wide coverage of commonly reported statistics (including t-tests, ANOVAs with 2 levels, and reported means), enabling consistent conversion across studies that vary in analytic approach. The main effects of two-way ANOVAs (between-subjects design) that feature 2 levels (df =1), and were not moderated by a significant interaction, were first converted using the *F*-to-*d* conversion (Thalheimer & Cook, 2020) and subsequently converted to *g*. Only unmoderated between-group main effects (df₁ = 1) were eligible for extraction. Effects that appeared only as simple effects or follow-up contrasts within a significant interaction were excluded (Borenstein et al., 2009). Additionally, studies reporting correlations, regression models, or general linear modeling were excluded because the reported statistics could not be converted to Hedges’. Neuroimaging papers or modeling papers were included *if* they provided behavioral measures that could be converted to Hedges’ *g*. For studies using prospect-theory models, we extracted the loss-aversion parameter (λ) rather than the utility-curvature risk parameter (γ/ρ), since λ reflects valence-sensitive decision processes consistent with the constructs analyzed across the meta-analysis. For a small subset that reported only binary choice proportions, g was computed from group proportions using esc_bin_prop(). When studies did not report convertible statistics, effect sizes were derived from figures using a plot-digitization tool, provided the figure contained adequate information (e.g., clearly identifiable means and error bars). Author contact was attempted when feasible, but response rates were very low, and none yielded usable data; therefore, figure-based extraction served as the primary method for recovering missing statistics. See PRISMA, Figure 2.

The PRISMA flow diagram reports the number of abstracts excluded based on the four inclusion criteria. Exclusion categories were assigned hierarchically. For example, if a study failed both the healthy-control criterion and the age criterion, it was categorized under “Unsuitable or missing healthy control group.” Email contact with authors was attempted during data extraction when necessary for missing information; however, no additional usable data were obtained, and these attempts did not affect screening or eligibility decisions. Additional exclusion reasons not shown in the PRISMA diagram include articles that were not in English, not peer-reviewed (e.g., dissertations), inaccessible full texts, or studies for which effect sizes could not be computed.

### Operationalization

Cognitive flexibility is the ability to adapt behavior in response to changing rules, contingencies, or task demands. Therefore, we included studies using tasks that required shifting between mental sets, updating response strategies, and inhibiting previously learned responses (Miyake, 2000; Diamond, 2013; Crawford, 1998). Supplementary Tables 1-4 detail the tasks and outcome measures to index cognitive flexibility. For the Trail Making Test (TMT), some studies reported Part B completion time, while others reported B–A difference scores. Reported measures were included as provided.

Decision-making was operationalized as performance on tasks requiring choice under uncertainty or risk (Bechara et al., 2013; Lejuez et al., 2002), involving learning from probabilistic reward, integrating reward to guide future outcomes, and tasks incorporating social considerations (Rilling & Sanfey, 2011). Tasks indexing this construct included the Iowa Gambling Task (IGT), Balloon Analogue Risk Task (BART), probabilistic learning paradigms, effort-based decision-making tasks, social decision-making tasks (e.g. Ultimatum Game), and differential reward/punishment paradigms. Supplementary Tables 1-4 detail the task and outcome measures to index decision-making. For the IGT tasks, reported net scores were used whenever available to support comparability and strong operationalization. CANTAB tasks were only included if they specifically assessed decision-making as defined in this meta-analysis; no CANTAB decision-making tasks met this criterion, and all were excluded.

### Meta-analytic methods

Meta-analytic computation and forest plots were conducted in ‘*meta’* and *‘dmetar*.’ (Harrer et al. 2019). The characteristics of each study included in the meta-analysis are described (see Supplementary Table 1-4). Given that we anticipated a large amount of between-subject heterogeneity, we utilized a random-effects approach. In addition to accounting for sampling error, a random-effects approach can better account for heterogeneity in the population (e.g. outcome of interest may have been measured in different ways). The restricted maximum likelihood estimator (Viechtbauer, 2005) was used to calculate the heterogeneity variance ^2^

After computation of the effect size, the sign of the effect size was adjusted to aid in interpretability; negative effect sizes reflect worse outcomes for anxiety or depression relative to controls or control analogues. Pooled effect sizes were calculated using the inverse-variance method below (Equation 1). Equation 1 shows that the pooled effect equals the sum of the study-specific effect sizes weighted by their respective study weights, divided by the sum of those weights. The weight for each study is given as, 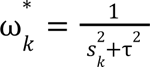

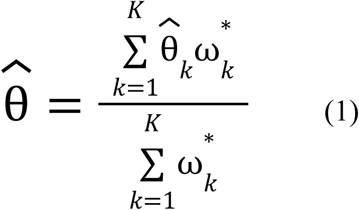

We used Knapp-Hartung adjustments (Knapp & Hartung, 2003) to calculate the confidence interval around the pooled effect. These adjustments have been shown to reduce the risk of a false positive (IntHout et al., 2014; Langan et al., 2019).

#### Outliers

Outliers were identified if the confidence interval of a given study did not overlap with the confidence interval of the pooled effect (Harrer et al. 2021).

## Results

### Heterogeneity results

Below we report estimates of between-subject heterogeneity for each outcome, using the frequently-reported metrics, τ^2^, I^2^, and Cochran’s *Q*. Although I^2^ is deemed easily interpretable (Harrer et al, 2021), the reader should observe I ^2^ is not a perfect measure of heterogeneity (Borenstein et al. 2017; Borenstein, 2023; and see Harrer et al. 2021). We provide several measures of heterogeneity for purposes of comprehensive reporting. In order to determine whether a pooled effect may be disproportionately impacted by the presence of outliers, the data-driven approach recommended by Harrer et al. (2021) was followed.

Specifically, the *Q* value and I^2^ value were inspected, and outlier analyses were conducted when moderate heterogeneity was detected (i.e. large Q values and I^2^ > 50.). While no singular value of *Q* or I^2^ has been proposed, these methods importantly avoid removing outliers without a stringent rationale (e.g. using a hunch or heuristic).

For the full model for anxiety on decision-making (k =22), the between-study heterogeneity variance was estimated at τ^2^ = 0.5013, 95% CI [0.2605; 1.2403], with an I^2^ = 81.0%, 95% CI [72.1%; 87.1%]. Examining the CI for τ^2^, the CI does not contain 0, suggesting heterogeneity exists. A *Q* test for heterogeneity showed significant heterogeneity, *Q(21)* = 110.53, *p* < 0.0001. Overall, results indicate high amounts of between-subject heterogeneity, which may suggest that the pooled effect may be influenced by a study underestimating or overestimating the true effect size. Therefore, outlier analyses (comparison of the confidence interval of a study relative to the confidence interval of the pooled effect; see Methods) were conducted. Adjusting for outliers, heterogeneity was reduced, I^2^= 18.5%, 95% CI [0.0%; 52.9%, *Q(18)* = 22.09, *p* = 0.228.

For the full model for anxiety on flexibility (k =10), the between-study heterogeneity variance was estimated at τ^2^ = 0.1354, 95% CI [0.0313; 1.0247] with an I^2^ = 64.7%%, 95% CI [50.2%; 85.9%]. Examining the CI for τ^2^, the CI does not contain 0, suggesting heterogeneity exists. A *Q* test for heterogeneity showed significant heterogeneity, *Q(9)* = 25.49, *p* = 0.0025. Therefore outlier analyses were conducted. Adjusting for outliers, heterogeneity was reduced, I^2^ = 29.8%, 95% CI [0.0%; 67.5%, *Q(8)* = 11.40, *p* = 0.1803.

For the full model for depression on decision-making (k=42), the between-study heterogeneity variance was estimated at τ^2^ = 0.1938, 95% CI [0.1141; 0.3850] with an I^2^ = 74.2% [65.2%; 80.9%]. Examining the CI for τ^2^, the CI does not contain 0, suggesting heterogeneity exists. A *Q* test for heterogeneity showed significant heterogeneity, *Q(41)* = 159.19 p < 0.0001. Therefore outlier analyses were conducted. Adjusting for outliers, heterogeneity was reduced, I^2^ = 57.9% [38.8%; 71.1%], *Q(34)* = 80.82, p < 0.0001.

For the full model of depression on cognitive flexibility(k= 26), the between-study heterogeneity variance was estimated at τ^2^ = 0.22 [0.11; 0.54], with an I^2^ = 77.3% [67.1%; 84.3%]. Examining the CI for τ^2^, the CI does not contain 0, suggesting heterogeneity exists. These results were supported by a *Q* test for heterogeneity, *Q(25)* = 109.97 p < 0.0001.

Overall, results indicate high amounts of between-subject heterogeneity, which may suggest that the pooled effect may be influenced by a study underestimating or overestimating the true effect size. Given the evidence for substantial heterogeneity, outlier analyses were conducted. Adjusting for outliers, heterogeneity was reduced, I^2^ = 37.7% [0.0%; 62.7%], *Q(21)* = 33.69, *p* = 0.0391.

### Meta-analytic results

Next, we report the pooled effect sizes for each outcome. Following recommendations by Harrer and colleagues (2021) and Borenstein (2023), the prediction interval is also reported to aid interpretation (see. IntHout et al., 2016). For anxiety and decision-making, the full random effects model showed a moderate to large and negative effect, *g* = −0.6507, 95% CI [−1.0055; −0.2958]. After removing outliers (*k* =3), the random-effects model showed a moderate and negative effect, *g* = −0.4472, 95% CI [−0.5961; −0.2982]. Prediction interval results indicate that future studies may demonstrate negative effects for anxiety on decision-making, *PI* [−0.5816; −0.3127]. Overall, results show anxiety impairs decision-making and these effects are moderate. See Figure 3.

**Figure 3.**
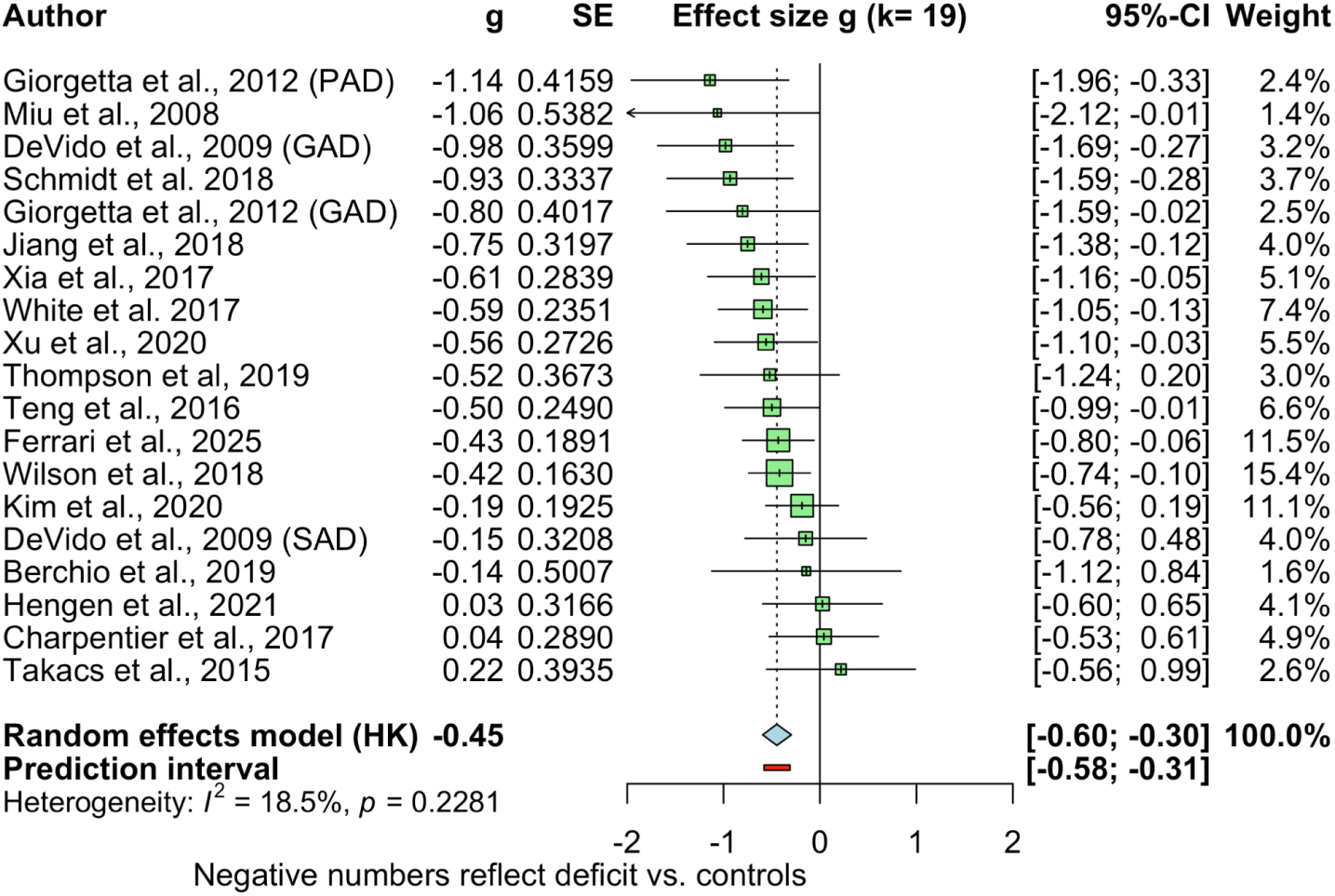
Pooled effects, Anxiety and decision-making. Negative effect sizes indicate worse decision-making outcomes for anxiety as compared to controls. Results indicate a moderate and negative effect, g = - 0.45. Plot shown with outliers removed (k =3). Disorder labels in parentheses indicate distinct samples within the same publication. Full citations are listed in the Appendix.

For anxiety and flexibility, a random effects model showed a small negative effect, *g* = −0.38, 95% CI: [−0.75; −0.008]. After removing outliers (k=1), a small negative effect was still observed, g = −0.25, 95% CI: [−0.48; −0.03]. The prediction intervals suggest that future studies may demonstrate negative effects for anxiety on flexibility, yet positive effects cannot be ruled out, prediction interval: *PI* [−0.68; 0.17]. Results therefore suggest that anxious disorders moderately impair flexibility. See Figure 4 for the meta-analytic results.

**Figure 4.**
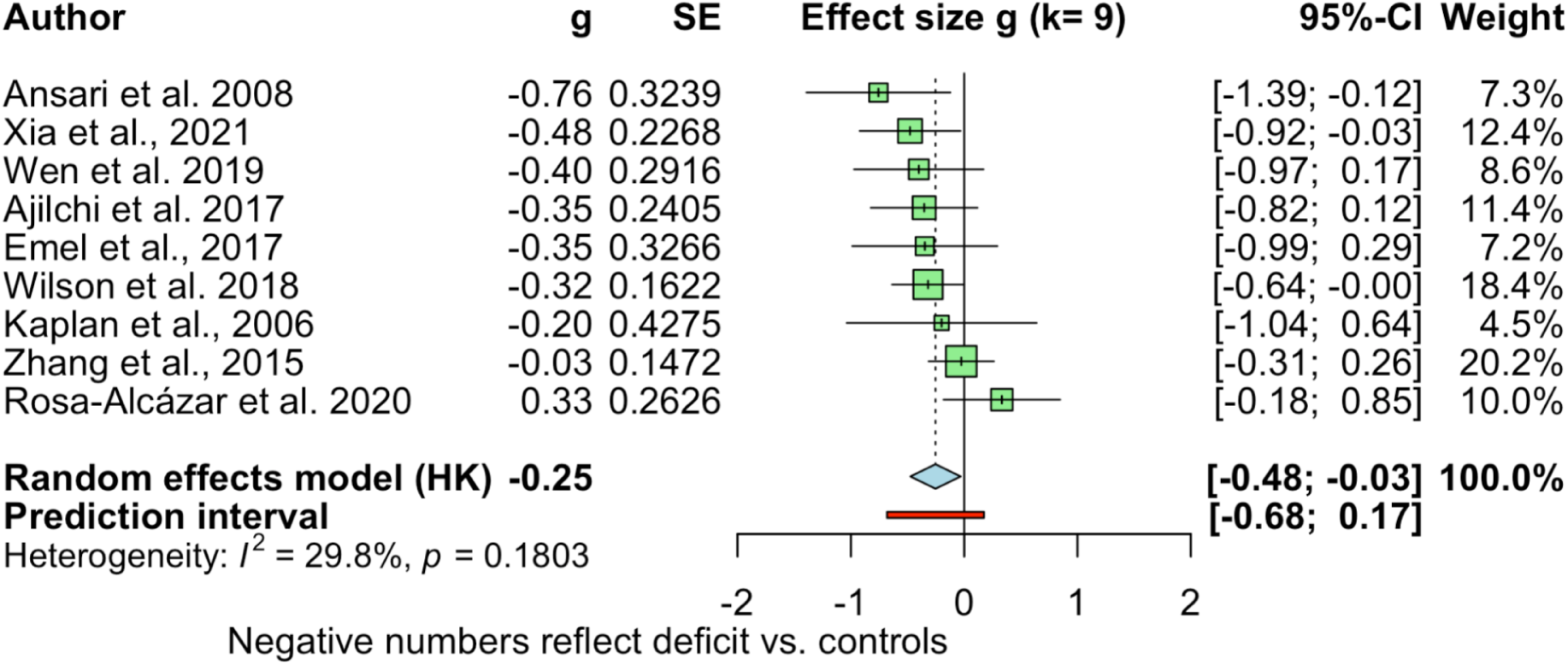
Pooled effects, Anxiety and flexibility. Negative effect sizes indicate worse flexibility outcomes for anxiety as compared to controls. Results indicate a moderate and negative effect, g = - 0.25. Plot shown with outliers removed (k =1).

For depression and decision-making, the random effects model showed small and negative effect, *g* = −0.3505, 95% CI [−0.5132; −0.1877]. After removing outliers (*k* =7), the random-effects model showed a moderate and negative effect, *g* = −0.3885, 95% CI [−0.5255; −0.2515]. Prediction interval results indicate that future studies may demonstrate negative effects for depression on decision-making but improved outcomes cannot be ruled out, prediction interval, *PI*= [−0.9995; 0.2225]. Overall, results show depression impairs decision-making but these effects are moderate. See Figure 5.

**Figure 5.**
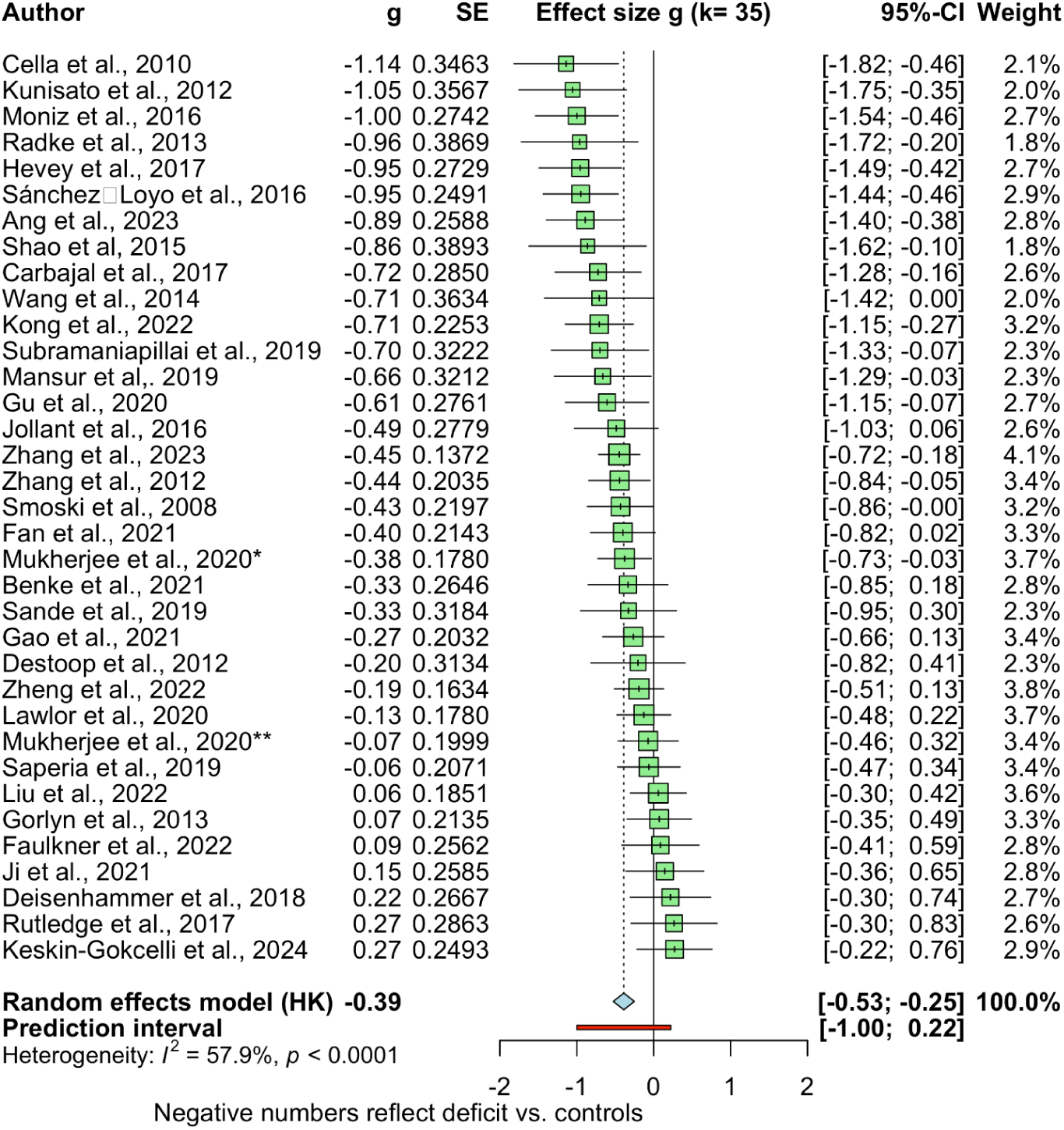
Pooled effects, depression and decision-making. Negative effect sizes indicate worse decision-making outcomes for depression as compared to controls. Results indicate a moderate and negative effect, g = - 0.3885. Plot shown with outliers removed (k =7). Note. * and ** indicate distinct studies by the same first author and year. Full citations for all studies are provided in the Appendix.

**Figure 6.**
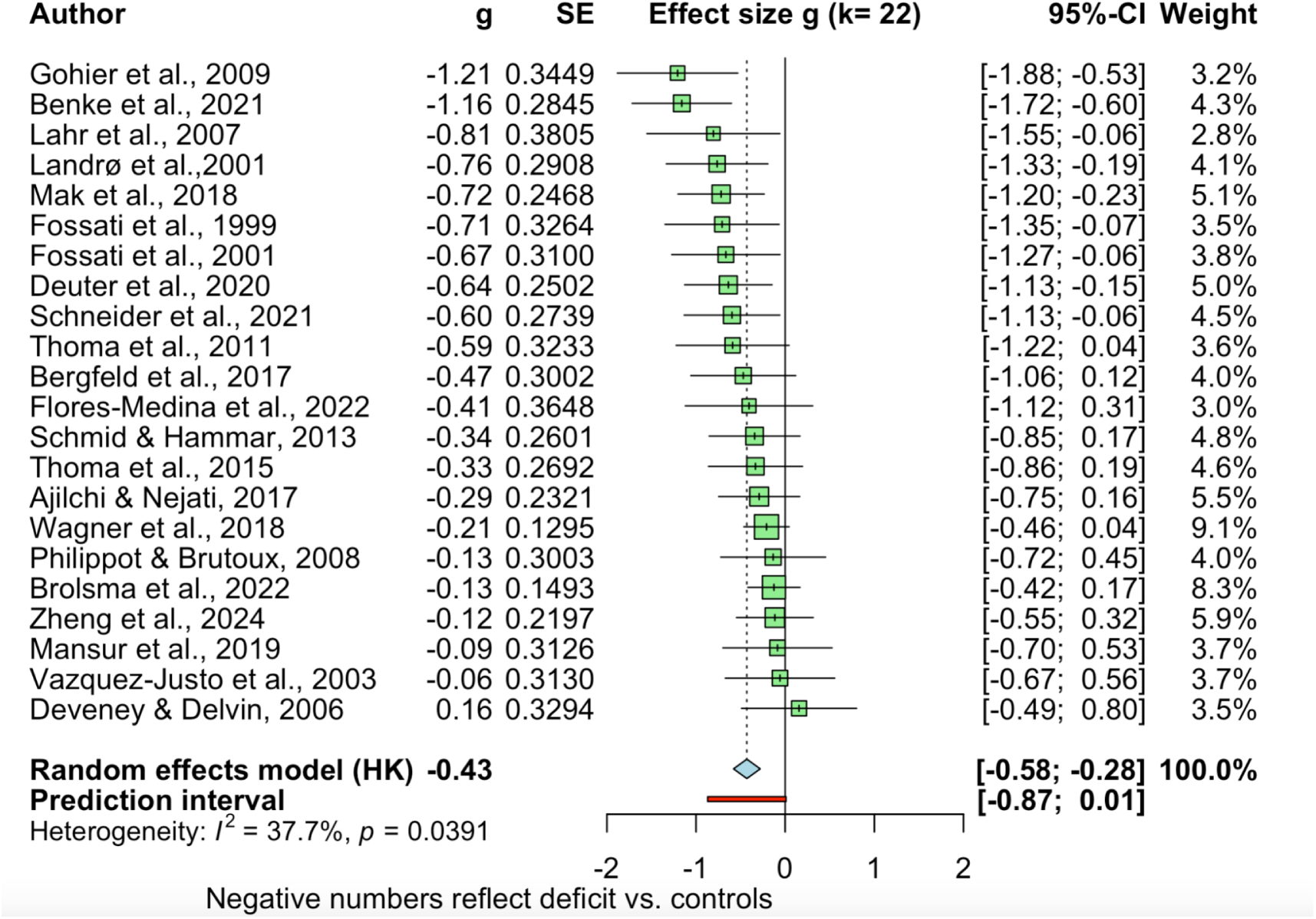
Pooled effects, depression and cognitive flexibility. Negative effect sizes indicate worse flexibility outcomes for depression as compared to controls. Results indicate a moderate and negative effect, g = - 0.43. Plot shown with outliers removed (k =4).

For depression and flexibility (k=26), the random effects model showed a moderate negative effect, *g* = −0.47, 95% CI: [−0.69; −0.24]. After removing outliers (*k* =4), the random-effects model showed a moderate negative effect, *g* = −0.4296, 95% CI: [−0.58; −0.28]. Prediction interval results indicate that future studies may demonstrate negative effects for depression on cognitive flexibility, prediction interval: [−0.87; 0.01]. Overall results suggest that depression impairs flexibility and these effects are moderate.

### Subgroup analysis

For decision-making outcomes, results did not support Hypothesis 1b: subgroup analyses indicated no significant differences between anxiety and depression in decision-making performance, *Q(1)* = 0.36 *p* = 0.548. Anxiety and depression show comparable impairments in decision-making, with no evidence that one group performs worse than the other. Both groups showed moderate impairments (anxiety: g = −0.45; depression: g = −0.39), with overlapping confidence intervals. See Table 2.

**Table 2.** Subgroup results for decision-making outcomes comparing depression and anxiety, outlier adjusted. Effect sizes are reported as Hedges’ g with corresponding 95% confidence intervals. Heterogeneity estimates (τ², τ, Q, and I²) are shown for each subgroup. The between-group test indicated no significant subgroup differences, Q(1) = 0.36, p = 0.548, suggesting comparable decision-making impairments across diagnostic categories.

| Subgroup | k | Hedges' g | 95% CI | $\tau^2$ | $\tau$ | Q (within) | I<sup>2</sup> |
| --- | --- | --- | --- | --- | --- | --- | --- |
| Anxiety | 19 | -0.4472 | [-0.5961, -0.2982] | <0.0001 | 0.0009 | 22.09 | 18.5% |
| Depression | 35 | -0.3885 | [-0.5255, -0.2515] | 0.0861 | 0.2934 | 80.82 | 57.9% |

| Subgroup Difference Test | Q | df | p-value |
| --- | --- | --- | --- |
| Between groups | 0.36 | 1 | .548 |

For flexibility outcomes, results did not support Hypothesis 2b. Subgroup analyses indicated no significant differences between depression and anxiety in flexibility, *Q*(1) = 2.15, *p* = .142. Both groups showed negative effects (depression: g = −0.43; anxiety: g = −0.25), with overlapping confidence intervals. See Table 3.

**Table 3.** Subgroup results for flexibility outcomes comparing depression and anxiety, outlier-adjusted. Effect sizes are reported as Hedges’ g with corresponding 95% confidence intervals. Heterogeneity estimates (τ², τ, Q, and I²) are shown for each subgroup. The between-group test indicated no significant subgroup differences, Q(1) = 2.15, p = .142, suggesting comparable flexibility impairments across diagnostic categories.

| Subgroup | k | Hedges' g | 95% CI | $\tau^2$ | $\tau$ | Q (within) | I<sup>2</sup> |
| --- | --- | --- | --- | --- | --- | --- | --- |
| Depression | 22 | -0.4296 | [-0.5794, -0.2798] | 0.0396 | 0.1990 | 33.69 | 37.7% |
| Anxiety | 9 | -0.2531 | [-0.4752, -0.0310] | 0.0250 | 0.1580 | 11.40 | 29.8% |

| Subgroup Difference Test | Q | df | p-value |
| --- | --- | --- | --- |
| Between groups | 2.15 | 1 | .1422 |

### Sensitivity Analysis

Finally, we examined small-study effects using Egger’s test to evaluate the robustness of the overall effect estimate. For anxiety and decision-making, Egger’s regression test indicated significant funnel plot asymmetry for the full model, β₀ = - 3.068, 95% CI [−5.88, −0.25], t = −2.135, *p* = 0.045. Although Egger’s test indicated funnel plot asymmetry, we did not conduct the frequently used trim-and-fill method because it is not considered reliable when the underlying meta-analytic model exhibits substantial heterogeneity (Peters et al., 2007; Terrin et al., 2003; Simonsohn et al., 2014), as was the case in the full model prior to outlier adjustment. Accordingly, the outlier-adjusted random-effects model remains the primary estimate. A funnel plot was constructed to visualize the distribution of study effects and precision. See Supplementary Figure 1.

For anxiety and flexibility, Egger’s regression test indicated no presence of funnel plot asymmetry in the full model, β₀ = - 2.46, 95% CI [−5.21, −0.29], t = −1.75, *p* = 0.118.

For depression and decision-making, Egger’s regression test indicated no presence of funnel plot asymmetry in the full model, β₀ = - 2.358, 95% CI [−4.71, - 0], t = −1.963, *p* = 0.0565.

For depression and flexibility, Egger’s regression test indicated no presence of funnel plot asymmetry in the full model, β₀ = - 0.866, 95% CI [−3.33, −1.6], t = -.688, *p* = 0.498.

## Discussion

### Summary

Research has yet to systematically review the influence of anxiety and depression on both decision-making and cognitive flexibility. The present study conducted a meta-analysis of anxious and depressive disorders on cognitive flexibility and decision-making outcomes. We used machine-learning tools to facilitate the screening of articles while maintaining human review, ensuring articles were properly vetted for meta-analytic criteria. Results demonstrate a negative effect across all outcomes, suggesting that both anxiety and depression are linked with impaired flexibility *and* decision-making. These meta-analytic results were then tested for subgroup differences. For decision-making outcomes, results showed no subgroup differences, *p* > 0.05, thus, failing to support our hypothesis that anxiety would demonstrate larger decision-making deficits as compared to depression. Similarly, for flexibility outcomes, we found no subgroup differences for anxiety and depression.

### Mechanisms implicated in poor decision-making

Both anxiety and depression were associated with impaired decision-making in the present study, consistent with known disruptions to key neurobiological substrates. Prior literature has established key neurobiological substrates that impact decision-making, including the amygdala and portions of the prefrontal cortex. When confronted with ambiguous choices or valence-based choices, amygdala activity drives computations towards threat-related signals, distorting how that reward information is weighted for optimal decision-making. With respect to anxiety, individuals with anxiety respond more strongly to ambiguous emotional face stimuli than individuals with less anxiety; a phenomenon said to be driven in part by hyper-activity of the amygdala, elevating both cognitive and affective responses to a threat stimulus (Hartley & Phelps, 2012b; Richards et al., 2002). Critically, damage to the amygdala impairs decision-making and judgments of social behavior (Bechara et al., 1999; Tranel & Hyman, 1990). Interestingly, known neural substrates of decision-making, such as the ventrolateral PFC, are also recruited in emotion and mood regulation (Mitchell, 2011), in part explaining why deficits in brain regions that regulate moods might directly alter decisions. Damage to the vmPFC impairs decision-making (Bechara, Damasio, Damasio, & Lee, 1999). Thus, portions of the PFC and the amygdala are prime candidates that could explain why both these disorders were associated with deficits in decision-making.

### Disorder-linked mechanisms in decision-making

Although the subgroup analysis showed no differences between anxiety and depression, the prediction interval evidence motivates future investigation into differences, but does not alter the null subgroup conclusion. The use of the prediction interval for interpreting non-significant pooled effects has been strongly recommended in previous literature (e.g. IntHout et al. 2016).

Specifically, for decision-making outcomes, the prediction intervals for *depression* showed a much wider prediction interval spanning zero, *PI*= [−0.9995; 0.2225], suggesting that future studies may show *improved* outcomes for those with depression. In contrast, prediction intervals for anxiety were overwhelmingly negative effects *PI* [−0.5816; −0.3127]. Notably, this pattern does not contradict the null subgroup finding but suggests that the two disorders may diverge in replication, with anxiety showing more consistent decision-making deficits across future research. Given these results, it is important to evaluate potential mechanisms that may differentially impact decision-making in anxious individuals as compared to depression. Prior literature has examined neural correlates that may partly explain why anxiety could be more strongly linked to decision-making deficits. For instance, the bed nucleus of the stria terminalis (BNST) is strongly implicated in both fear and anxiety (Davis et al. 2010),and linked to GAD (Duvarci et al. 2009; Avery et al. 2016; Avery et al. 2014). Given its role in sustained threat anticipation, BNST activity may bias decision-making under ambiguity toward threat-avoidant choices, particularly in GAD, where uncertainty intolerance is pronounced. Further, the orbitofrontal cortex, a region frequently cited in the context of decision-making, is said to aid in risky and ambiguous decisions (Krain et al., 2006). Critically, the orbitofrontal cortex has been implicated in anxiety disorders (Milad & Rauch, 2007). Beyond the OFC, the amygdala, once believed to be an anxiety-specific neural substrate for decision-making is now documented and studied in depression (see Grogan et al., 2022). Notably, studies show consistent amygdala hyperreactivity in MDD (Li & Wang, 2021; Janiri et al., 2020, but see Grogans et al., 2022).

Despite the link to depression, the amygdala is a candidate explanation for anxiety-related decision-making, given that the amygdala may also play a role in the generation of early anxious behavior. For instance, animal work suggests that behavioral transitions of low and high fear states in anxiety might be mediated by basolateral amygdala (Herry et al., 2008), whereas the mechanisms for depression are said to involve disruption of the nucleus accumbens to generate appetitive responses to arousing stimuli (see Lemos et al., 2012).

### Transdiagnostic factors implicated in decision-making

We next consider transdiagnostic factors that can explain why our meta-analytic results showed decision-making deficits in anxiety and in depression. Prior available theories that describe specificity for anxiety in decision-making have been recently challenged. For instance, theories for intolerance of uncertainty (IU), a well-documented factor in decision-making (Meeten et al., 2012; Pulcu & Browning, 2019), were previously believed to be uniquely characteristic of anxiety disorders (Norton et al., 2005; Sexton et al., 2003). Nevertheless, meta-analytic evidence has suggested that IU may also have a role in depression (see Gentes & Ruscio, 2011). Research therefore suggests that IU is a transdiagnostic factor that contributes to multiple psychopathological outcomes, including depression. Indeed, there is growing recognition that anxiety and depression share transdiagnostic factors once thought to be anxiety-specific or depression-specific. Taken together, future studies should test these factors as potential mediators that might explain why anxiety and depression equally demonstrate worse decision-making.

### Flexibility Outcomes Show Comparable Impairments in Anxiety and Depression

Turning to our flexibility results, the present study showed that both anxiety and depression demonstrate impaired cognitive flexibility. The subgroup analysis indicated no significant differences between the two disorders. The pattern in these results reflects shared disruptions in cognitive control processes, rather than disorder-specific mechanisms. Across set-shifting and task-switching paradigms, individuals across clinical or trait-level anxiety or depression showed difficulty updating task rules, adapting behavior in response to contingencies, or disengaging from prior mental sets.

Several candidate mechanisms may contribute to these shared impairments. We first evaluate prior well-established candidate explanations. For instance, although negative affect is a core feature of depression, the underlying mechanism linking affective valence to flexibility performance is poorly understood(van Dooren et al., 2021). Additionally, there is mixed support for linking affect to flexibility performance (Ashby et al., 1999; Grol & De Raedt, 2021, Hsieh & Lin, 2019; Liu & Wang, 2014). High comorbidity between anxiety and depression further complicates interpretation, as interactive effects have been reported in some studies (Dotson, 2014; Yoon & Joormann, 2012, Wu et al. 2016), but not others (O’Shea et al. 2016). Taken together, these inconsistencies suggest that neither negative affect nor comorbidity alone can account for the flexibility deficits observed across disorders. In the present study, the most frequent anxiety disorder falls under a distress-based (e.g. GAD and STAI). The shared overlap between distress-based anxiety and MDD may help explain the comparable deficits observed across anxiety and depression in the present study.

### Working Memory Capacity and Perseverative Negative Thought

Another commonly investigated moderator of flexibility outcomes in clinical populations is working memory capacity. Cognitive flexibility paradigms, such as task-switching, place a substantial load on working memory (i.e. on a difficult task), which can impede the individual’s ability to inhibit distractors (Berggren & Derakshan, 2013; Gazzaley, 2011). In laboratory settings, for example, the manner in which stimuli are shown can impact the load on working memory (Wimmer & Poldrack, 2022). Nevertheless, both anxiety and depression are linked to disruption in working memory. For example, depressive symptoms are associated with poor short-term working memory; individuals with the highest depression scores show poorer performance on very short-term (1-back) representation maintenance (Dotson, 2014). Similarly, poor working memory capacity has been shown in high-trait anxiety (Moran, 2016). Given the pattern in our flexibility results, working memory deficits do not fully account for the results in the present study. We had hypothesized that depression would show larger flexibility deficits, given that a long-established component of depression is rumination, which involves repetitive negative thoughts about past experiences. The explanatory power of this argument is weakened given that similar repetitive negative thinking was recently observed in other anxiety disorders (Olatunji et al., 2013), challenging previous traditional characterizations. For instance, Shihata and colleagues (2022) recently noted that frequently used measures of perseverative negative thinking, including the Penn State Worry Questionnaire and the Ruminative Response Scale, are measuring trait-level tendencies for repetitive negative thinking across both anxious and depressive disorders, including GAD, SAD, and depression. Taken together, the evidence suggests that rumination alone cannot wholly account for the shared flexibility deficits we report.

### Transdiagnostic factors for general cognitive outcomes

Anxiety and depression are characterized by their shared neurobiological complexity (see LeDuke et al., 2023). The findings of the present study demonstrate that anxiety and depression are associated with deficits in both decision-making and cognitive flexibility. Our meta-analysis results reflect outcomes across several task measures. Because these impairments emerged across multiple paradigms and did not differ between disorders, the pattern is consistent with shared transdiagnostic mechanisms rather than disorder-specific effects. We note, however, that comparable effect sizes across disorders do not rule out disorder-specific mechanisms that could independently converge on similar performance deficits. For instance, anxiety may impair decision-making primarily through threat-biased valuation and intolerance of uncertainty, while depression may do so through blunted reward sensitivity and anhedonia. Future studies employing computational modeling approaches may be able to distinguish between transdiagnostic and disorder-specific factors including learning rates, loss aversion, and reward sensitivity, thereby bringing further insight into our meta-analytic evidence of comparable deficits.

Transdiagnostic research and the transdiagnostic approach (Krueger & Eaton, 2015) suggest that multiple factors cut across a variety of pathology. We now review advances from the transdiagnostic literature and highlight three factors influencing cognitive outcomes: Neuroendocrine, environmental, and neurobiological factors. First, the interaction of neuroendocrine factors and environmental factors cuts across both anxiety and depression.

Notably, stress and adversity are predictors of both anxiety and depression. Rodent studies show that early adversity leads to anxiety via generalized fear of non-predictive cues and disrupted prediction error signaling (Wright et al., 2015). Severe and chronic stress also induces anxiety-like behavior (Pohl et al., 2007). Further work has probed the role of the central amygdala in releasing corticotropin releasing factor (CRF), a neuroendocrine that signals in the hypothalamic pituitary adrenal axis; notably, CRF is a biomarker in both anxiety and depression (Hauger et al., 2009). Notably, these findings underscore how neuroendocrine and environmental factors can generate vulnerabilities across disorders.

Next, recent neuroimaging efforts utilizing a transdiagnostic approach have advanced our understanding of shared psychopathology and cognitive impairment. For instance, a recent meta-analysis showed that a large swath of psychotic and non-psychotic disorders, including depression and anxiety, were associated with gray matter loss in the anterior insula and dorsal anterior cingulate (dACC) (Goodkind et al., 2015; McTeague et al., 2016). Notably, these regions have been implicated in decision-making (Smith et al., 2014) and conflict-monitoring. Further, recent reviews have suggested a shared *cognitive dyscontrol that* cuts across psychopathology including psychotic, bipolar, depression, anxiety, and substance use disorders (McTeague et al., 2016). Taken together, converging transdiagnostic evidence from neuroendocrine and neurobiological research indicates that anxiety and depression are similarly linked to cognitive impairment, providing a probable explanation for the comparable deficits observed in the present meta-analysis. This transdiagnostic perspective is well-situated within a broader framework of psychopathology. Supporting this view, this evidence aligns with a *biopsychosocial* model (Engel, 1980), in which anxiety and depression arise from complex interactions among biological, psychological, and social factors (Buckner et al., 2013; Garcia-Toro & Aguirre, 2007; Taschereau-Dumouchel et al., 2022). Moreover, our results are consistent with dimensional models of psychopathology, such as the RDoC framework or integrations of RDoC with HiTOP (e.g. Michelini et al., 2021), which emphasize cross-cutting cognitive and affective processes rather than disorder-specific categories. Recent work has sought to reconceptualize once-believed discrete disorders under the perspective of HiTOP and RDoC (e.g. Hawn et al. 2022).

### Machine-learning in Meta-analytic Workflows

Meta-analyses are complex studies that involve substantial logistical planning. An additional strength of the present study was the use of machine-learning tools to support the initial literature screening process. In particular, the cognitive literature on anxiety and depression is vast and heterogeneous, spanning tens of thousands of records across multiple task paradigms and reporting conventions. Manual screening alone would have been prohibitively time-consuming and risked inconsistent coverage. Machine-learning–assisted screening is an emerging and effective approach for managing this scale of literature, and in the present study, it enabled efficient identification of potentially relevant articles. All machine-flagged records were subsequently hand-verified to ensure alignment with meta-analytic criteria and to extract compatible statistics. This workflow also highlights how machine-learning–assisted screening can support future large-scale cognitive meta-analyses, particularly in domains with heterogeneous task structures.

### Future directions

#### Re-thinking disorder-specific approaches

The present study reflects an important step for examining the impact of anxiety and depression on task-based cognitive outcomes via meta-analytic evidence. One important implication of the present results is that it may potentially inform a *staged model f*or treating anxiety and depression; staged models (classifying disease progress in several stages) provide health care professionals with key insight into the development of a disease and the most appropriate treatment during that stage. Indeed, there is growing progress on this front, as researchers advocate moving away from binary classification of depression to a staged model that can account for severity and disease development (Patel, 2017). Notably, Fried (2015) highlights the substantial heterogeneity within depression, showing that individuals can present with very different symptom profiles despite sharing the same diagnosis. This heterogeneity limits the usefulness of categorical diagnostic systems and strengthens the rationale for staged or dimensional models that capture symptom severity and progression. Fried’s recent work on symptom-level heterogeneity and dimensional, cluster-based models underscores why categorical diagnoses fall short and why staged approaches are increasingly needed.

Significant gaps remain in our understanding of affective pathology on behavioral outcomes. One of the most frequent approaches (disorder-specific approach) involves investigating the role of each disorder’s contribution on some measure of executive function (e.g. cognitive flexibility, working memory). For instance, the finding that individuals with severe MDD show pronounced executive function impairments (Steele et al. 2007), reflects a disorder-specific approach. Unfortunately, the disorder-specific approach is limited by incorrect underlying assumptions about the discreteness of a given disorder. Research has proposed hypotheses describing how transdiagnostic factors impact cognitive outcomes. For instance, in scenarios that require cognitive control, factors that cut across anxiety and depression influence the neural computation underlying the expected value of control (EVC; Shenhav et al., 2013, 2016). Looking at a task-switching scenario, the cost-benefit calculus for a switch has been posited to be informed by the reward expectancy. Specifically, recent work has provided insight into the neural calculus that determines the amount of cognitive effort required, which is said to reflect an individual’s control allocation (Frömer et al., 2021). It is therefore likely that transdiagnostic factors, including rumination and attentional control (Hsu et al., 2015), may indirectly or directly disrupt an individual’s ability to determine the amount of cognitive effort required. Thus, tasks that require cognitive control may show comparable deficits in both anxiety and depression because a poor calculus is made for the cognitive effort required.

#### HiTOP predictions

The recent taxonomy HiTOP (Kotov et al., 2017) demonstrates that the surrounding psychopathology, including anxious and depressive disorders, may be better explained by subfactors shared across disorders. Relevant to the present study, HiTOP classifies GAD and MDD under a subfactor ‘distress-based’ whereas other anxiety disorders like PAD and SAD are classified under a ‘fear-based’ sub-factor. Despite the factor-analytic and meta-analytic studies (e.g. Ringwald et al., 2023) in support of HiTOP, the DSM and ICD remain the prevailing framework for psychopathology; future research should test the predictions of the HiTOP model. One notable and recent example was the meta-analysis by Ringwald and colleagues (2023). The present studies were analyzed by the reported clinical diagnosis (e.g. GAD) or trait-level (trait anxiety). In the present study, we were unable to test the predictions of HiTOP given an imbalance of studies to test for differences. In order to more accurately compare effect sizes, future studies need to address this striking imbalance in the literature for depression studies; for instance, a pertinent review (Castaneda et al., 2008) mentioned that the overwhelming majority of studies examining depression on cognitive impairment are primarily individuals with MDD, leaving other depressive disorders (e.g. dysthymia) unaccounted for. In the present study, the majority of studies included for depression were of an MDD diagnosis.

Overall, our findings add to a growing body of meta-analytic work (e.g., Buur et al., 2025; Hong & Cheung, 2015) supporting a transdiagnostic perspective on across and anxiety-and mood-related psychopathology. Beyond reinforcing this broader framework, the present results also offer insight into how task paradigms may continue to evolve to more precisely capture the underlying construct. Importantly, because our meta-analysis included both clinical and trait-level anxiety and depression, the findings speak to the severity and continuity of decision-making deficits across the full spectrum of anxiety and depression pathology.

### Limitations

Decision-making is a multifaceted construct; the present study examined diverse aspects of decision-making, including decisions under risk, advantageous choice selections and social decision-making. A limitation of the present study is the underrepresentation of depression in contexts of social decision-making, given that few studies have evaluated this outcome in individuals with depression. This scarcity is notable considering that depression symptoms are associated with poor reward learning in social contexts (Safra et al., 2019). Turning to the sensitivity analyses, for depression and decision-making there was an indication of no significant funnel plot asymmetry (β₀ = −2.358, *p* = .0565), whereas for anxiety and decision-making, Egger’s test indicated significant asymmetry (β₀ = −3.068, *p* = .045). Given the substantial heterogeneity and diversity of paradigms within the decision-making domain, this asymmetry likely reflects small-study effects driven by paradigm variability rather than selective publication. One possible explanation for asymmetry in anxiety but not depression is that anxiety research often involves subtype-specific investigations paired with highly tailored tasks. Finally, Egger’s tests for anxiety-flexibility and depression-flexibility outcomes were non-significant, suggesting funnel plot symmetry for flexibility outcomes.

Moreover, we note a greater representation of depression studies on flexibility outcomes as compared to anxiety studies on the same outcomes. Although the subgroup analysis indicated no significant differences between depression and anxiety in flexibility outcomes, *Q*(1) = 2.15, *p* = .142, the comparison should be interpreted cautiously, given the unequal representation of studies with depression contributing more than twice as many studies (*k* = 22) as anxiety (*k* = 9). This comparison may have been underpowered to detect a true difference, likely reflecting the broader underrepresentation of anxiety flexibility studies in the available literature. We suggest that future work should continue to investigate anxiety-related flexibility deficits; the results of the present study reflect the available evidence. Like past meta-analytic work (e.g. Buur et al., 2025; Hong & Cheung, 2015), our results should be interpreted cautiously given the large amount of between-subject heterogeneity (see Results). Further, the present study excluded studies with individuals with a comorbid condition to control for confounding effects. We observe this as a limitation given that individuals in the real-world often present with comorbid anxiety and depression as compared to either one alone (Merikangas et al., 2003), affecting the study’s generalizability. Overall, the meta-analytic results of the present study should be interpreted as an analysis of the publicly available effects across the wider cognitive domain.

#### Trait anxiety vs. State anxiety

The present meta-analysis excluded state-based anxiety effects found in the literature, given that state-based assessments (e.g. state-induced anxiety or state-based questions on the STAI self-report) capture how an individual feels in a specific moment, rather than an anxious trait. Future research should examine whether the present results can be extended to state-based anxiety measures. A limitation of meta-analysis is that the results are influenced by the publicly available literature; where possible, efforts were made to contact authors and request the appropriate data. We received a reply from one author but the provided data was not suitable for inclusion.

#### Self-report vs. Task-based measures

Important lines of research have considered the distinctions between self-report-based measures and task-based measures. First, it is recognized that cognitive task-based measures and cognitive self-report measures are capturing related but distinct constructs (Duckworth & Kern, 2011; Snyder et al., 2021; Toplak et al., 2013). A key advantage of task-based cognitive measures is that they avoid social desirability biases linked to self-report measures (Dang et al., 2020; Hohl & Dolcos, 2024; Howlett et al., 2021); task-based measures are also regarded for their structured approach to capture largely autonomic responses (Hohl & Dolcos, 2024; Howlett et al., 2021). Given the advantages of task-based measures, the present study excluded articles that used self-report measures. Future studies should examine if distinct constructs that are captured by self-report measures also extend to the measures captured in a task.

## Conclusion

The present study conducted a meta-analysis to examine the effects of anxiety and depression on decision-making and cognitive flexibility. Overall, both disorders were associated with deficits in these domains, with small to moderate negative effects for both decision-making and flexibility. Across outcomes, we found no significant differences between anxiety and depression, consistent with a transdiagnostic perspective. Although anxiety and depression show distinct behavioral phenotypes, they also share underlying features that may contribute to comparable impairments in higher-order cognition. Across multiple paradigms and outcome measures, the present findings link both disorders to broad disruptions in decision-making and cognitive flexibility. Continued meta-analytic and empirical work is needed to clarify how these disorders shape human behavior and cognitive functioning.

## Data Availability

All data analyzed during the present study are contained within the manuscript (including Supplementary Tables and Figures).

## Supplementary Tables

**Supplementary Table 1.**
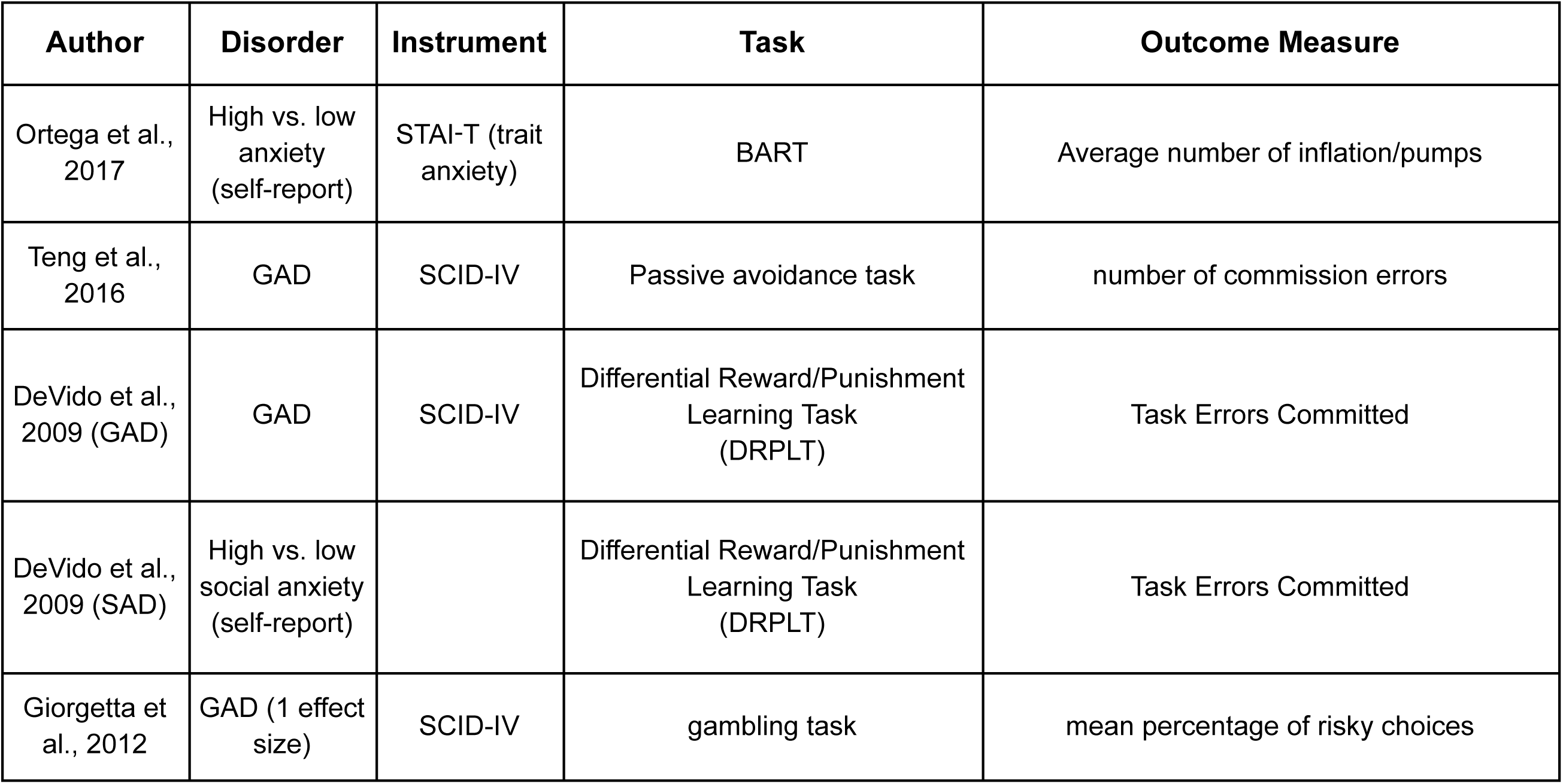
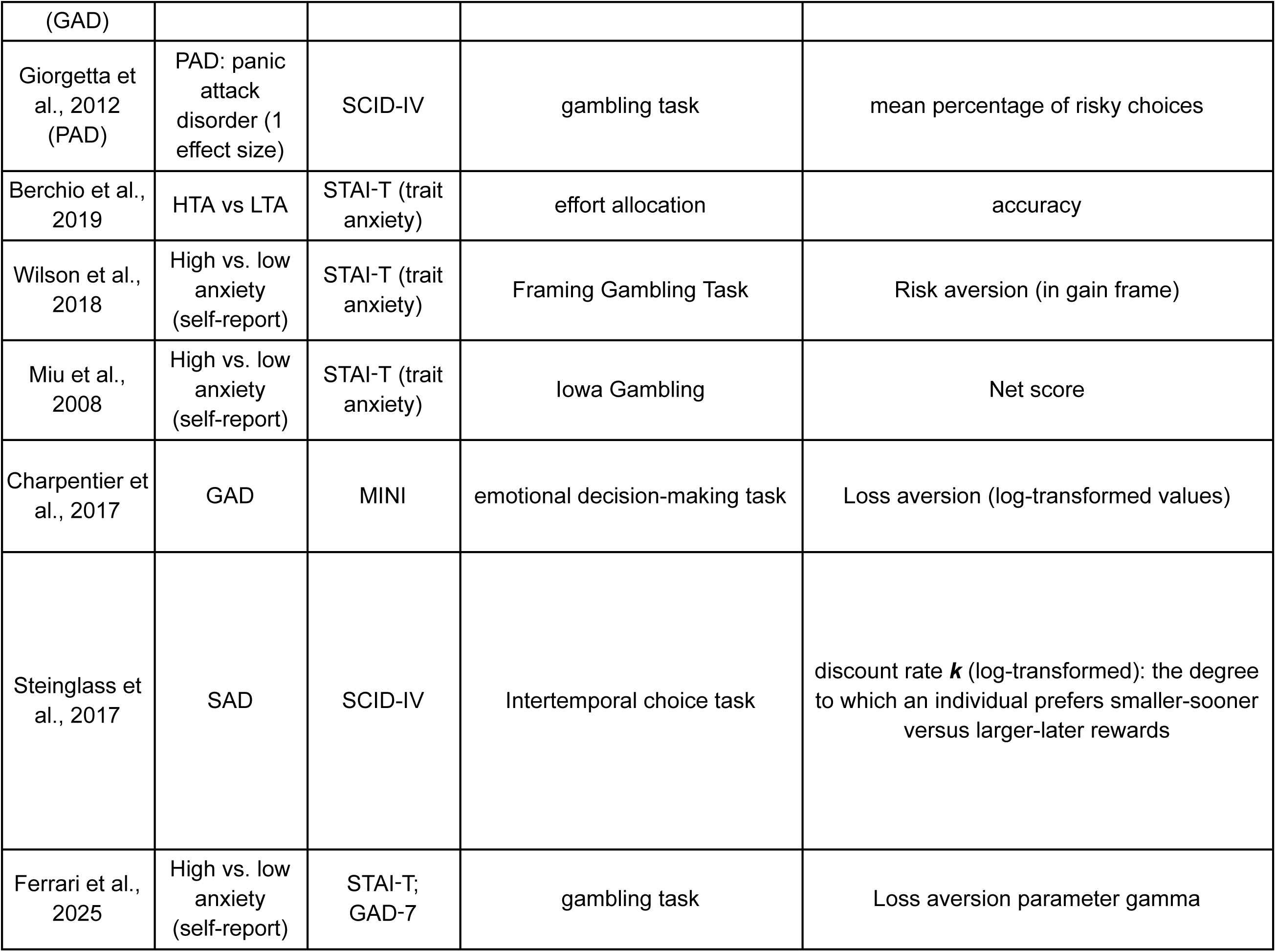
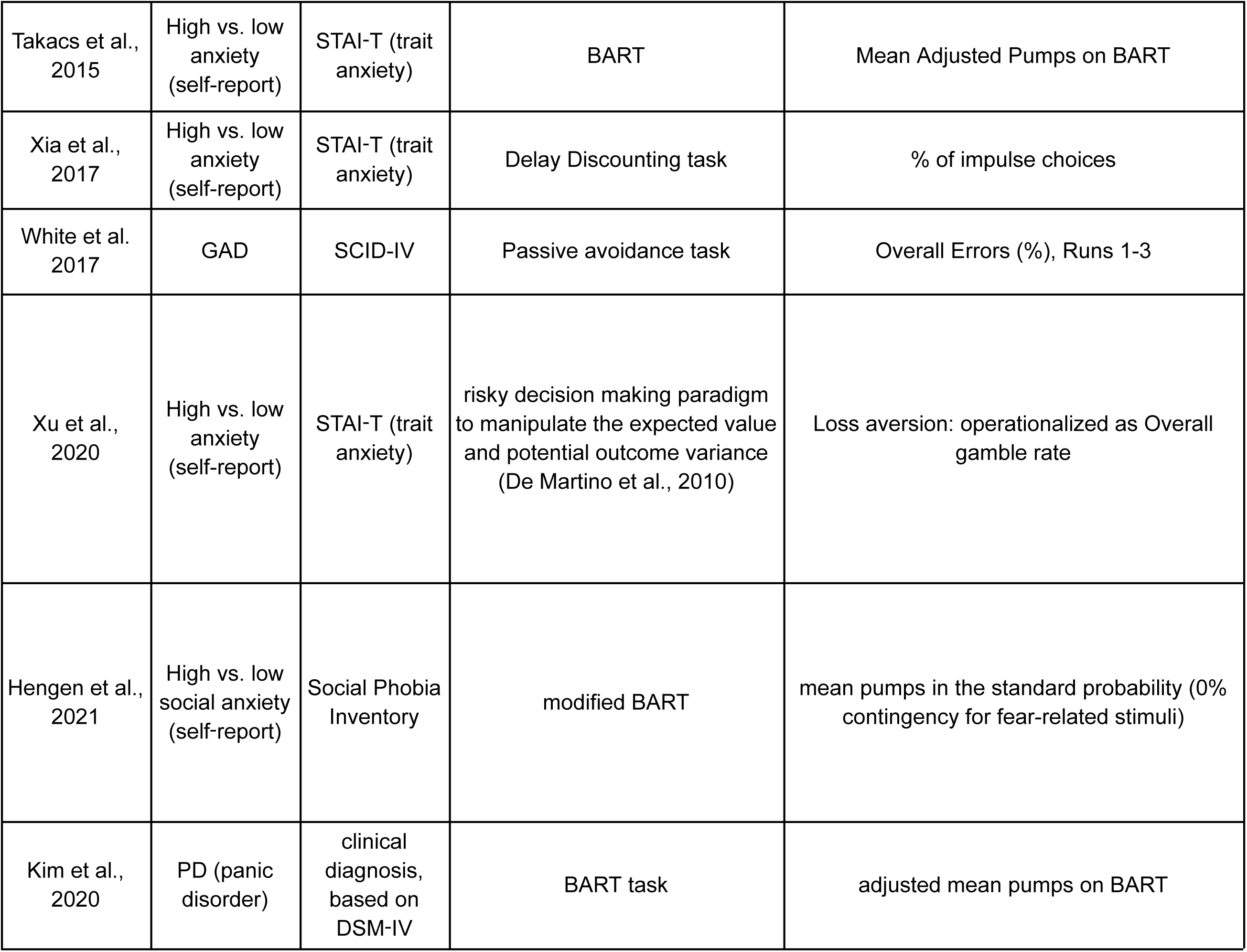
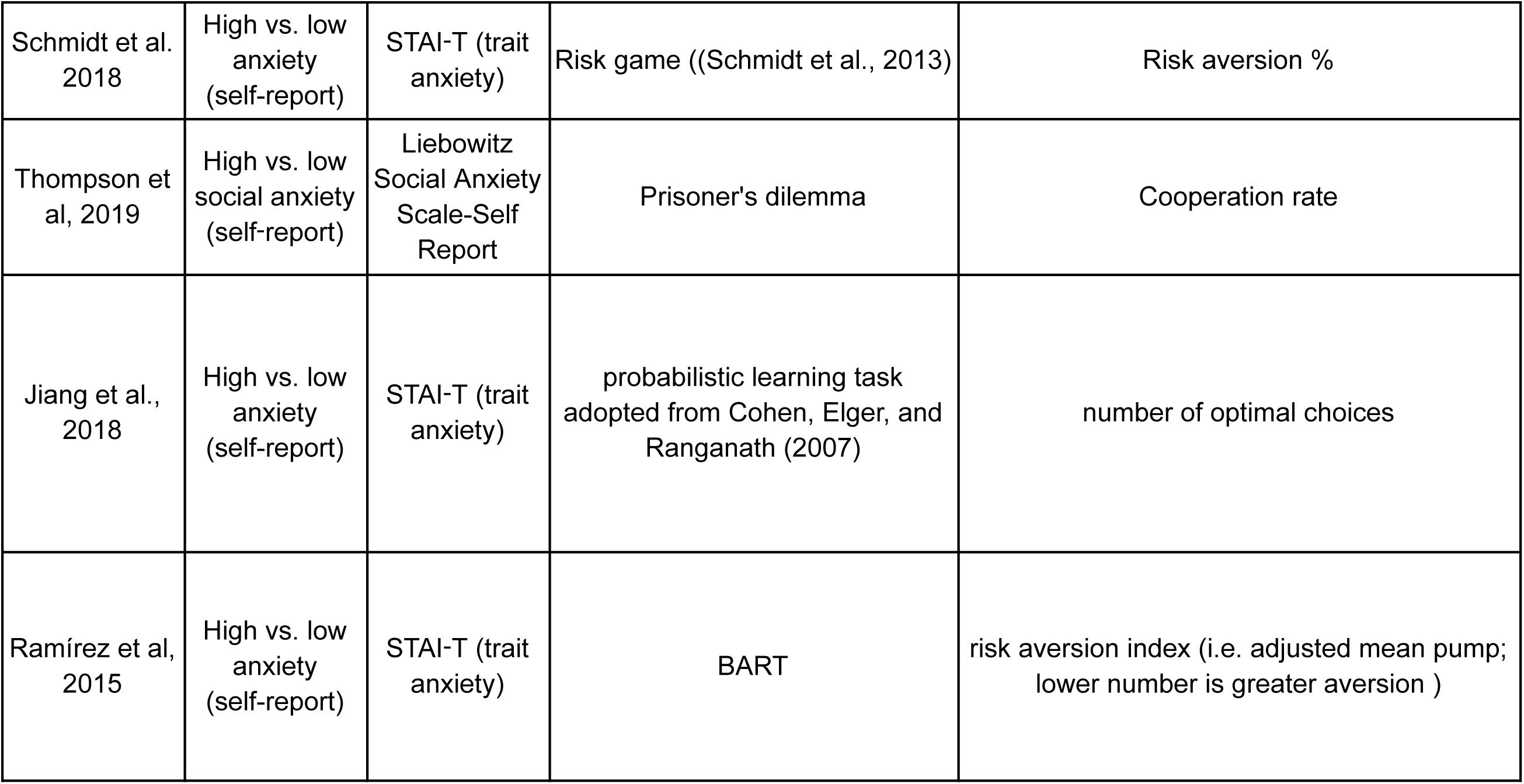
Effects of Anxiety on Decision-making: Summary of the characteristics of articles included in the meta-analysis for the full model ( includes articles that were established as outliers). Abbreviations: SCID= Structured Clinical Interview for DSM, PAD = panic anxiety disorder, GAD = generalized anxiety disorder, SAD = social anxiety disorder, STAI = State-Trait Anxiety inventory, ADIS = Anxiety disorder interview schedule, IGT = Iowa Gambling Task, BART = Balloon Analogue Risk Taking.

**Supplementary Table 2.**
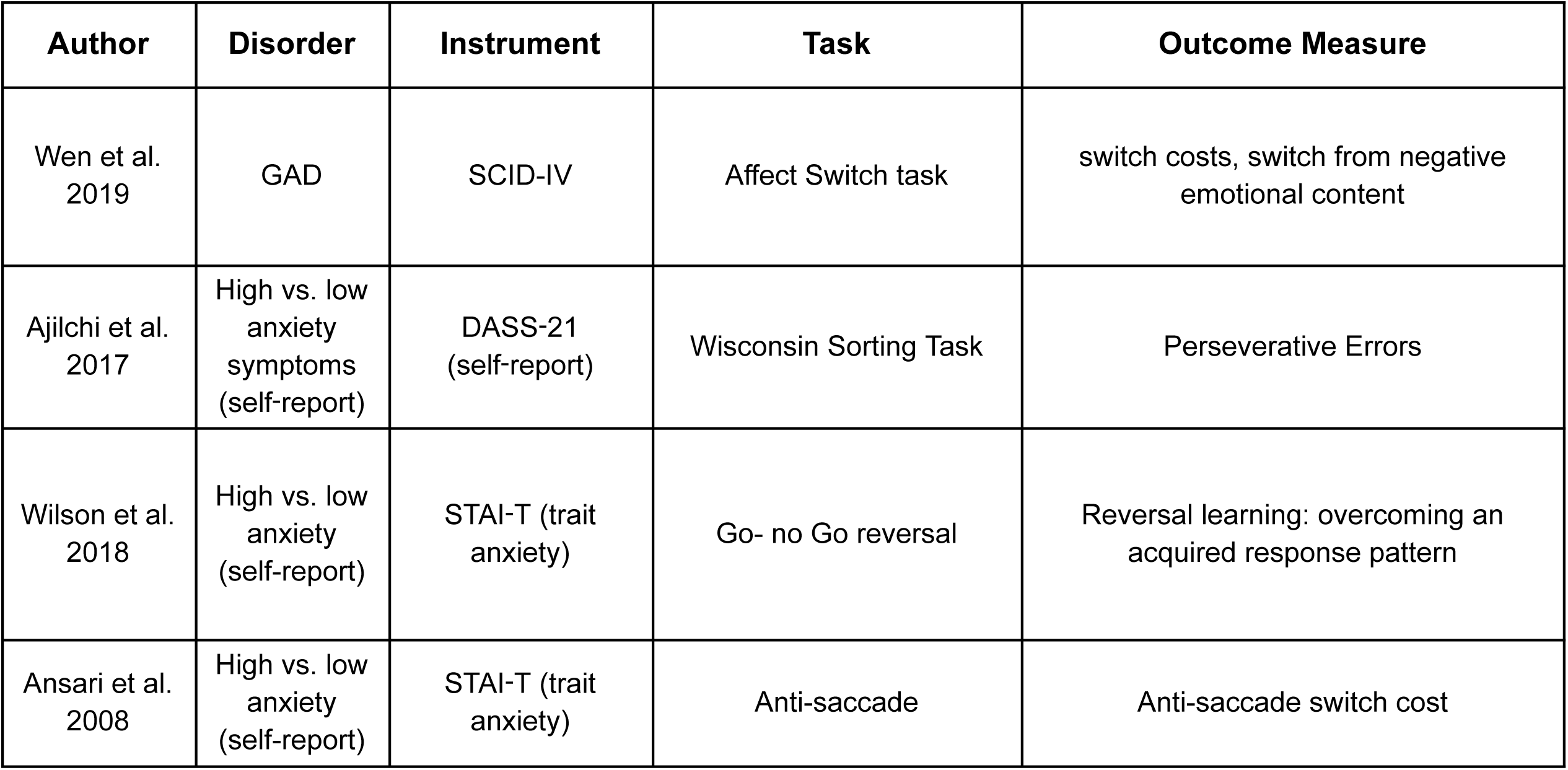
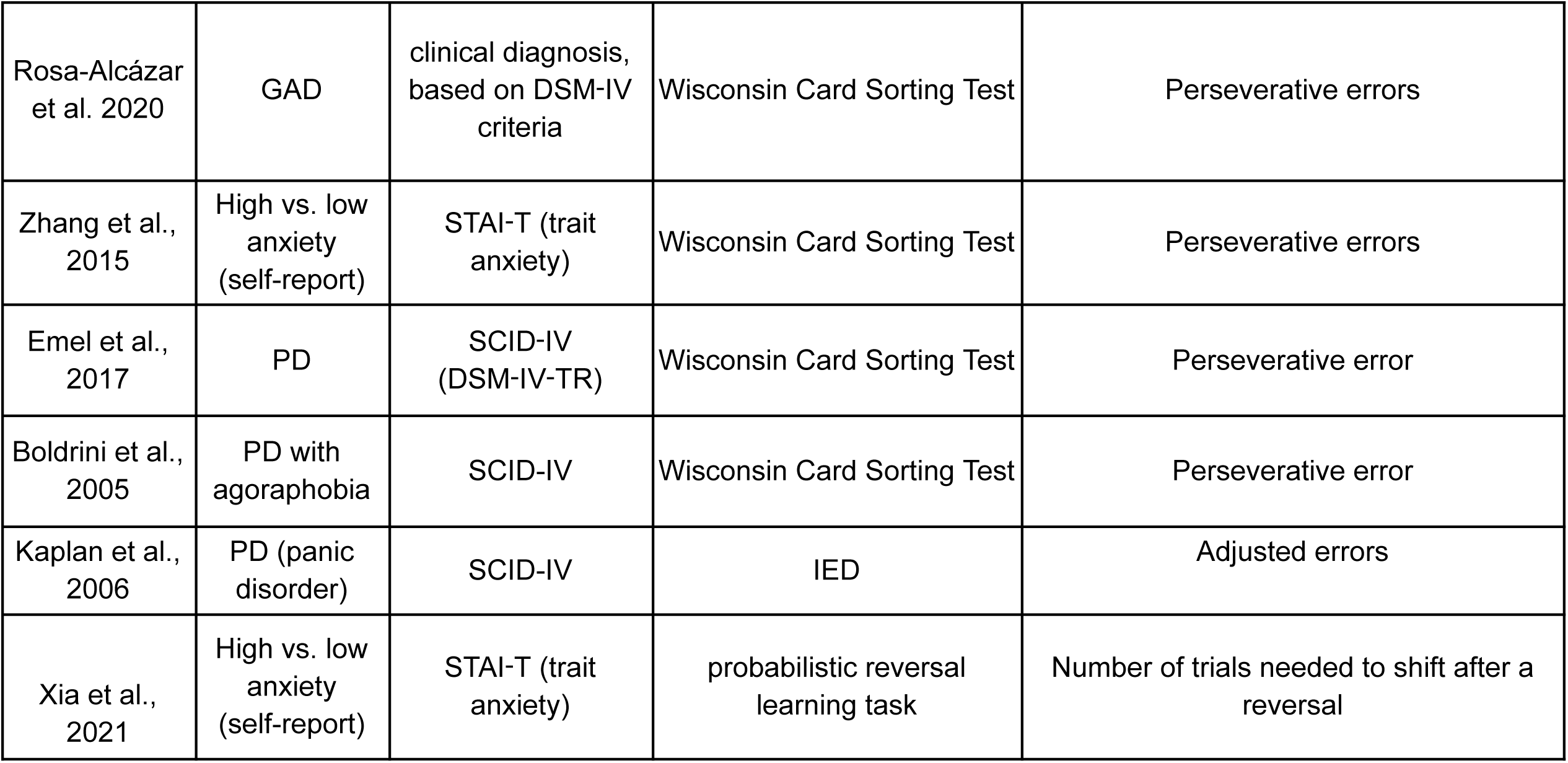
Effects of Anxiety on Cognitive Flexibility: Summary of the characteristics of articles included in the meta-analysis for the full model ( includes articles that were established as outliers). Abbreviations: GAD = generalized anxiety disorder, STAI = State-Trait Anxiety inventory, SCID = Structured Clinical interview, DASS = Depression, Anxiety and Stress Scale.

| Author | Disorder | Instrument | Task | Outcome Measure |
| --- | --- | --- | --- | --- |
| Wen et al.<br>2019 | GAD | SCID-IV | Affect Switch task | switch costs, switch from negative emotional content |
| Ajlchi et al.<br>2017 | High vs. low anxiety symptoms (self-report) | DASS-21 (self-report) | Wisconsin Sorting Task | Perseverative Errors |
| Wilson et al.<br>2018 | High vs. low anxiety (self-report) | STAI-T (trait anxiety) | Go- no Go reversal | Reversal learning: overcoming an acquired response pattern |
| Ansari et al.<br>2008 | High vs. low anxiety (self-report) | STAI-T (trait anxiety) | Anti-saccade | Anti-saccade switch cost |
| Rosa-Alcázar et al. 2020 | GAD | clinical diagnosis, based on DSM-IV criteria | Wisconsin Card Sorting Test | Perseverative errors |
| Zhang et al., 2015 | High vs. low anxiety (self-report) | STAI-T (trait anxiety) | Wisconsin Card Sorting Test | Perseverative errors |
| Emel et al., 2017 | PD | SCID-IV (DSM-IV-TR) | Wisconsin Card Sorting Test | Perseverative error |
| Boldrini et al., 2005 | PD with agoraphobia | SCID-IV | Wisconsin Card Sorting Test | Perseverative error |
| Kaplan et al., 2006 | PD (panic disorder) | SCID-IV | IED | Adjusted errors |
| Xia et al., 2021 | High vs. low anxiety (self-report) | STAI-T (trait anxiety) | probabilistic reversal learning task | Number of trials needed to shift after a reversal |

**Supplementary Table 3.** Effects of Depression on Decision-making: Summary of the characteristics of articles included in the meta-analysis for the full model ( includes articles that were established as outliers). HAMD = Hamilton Depression Scale, IGT = Iowa Gambling Task, BART = Balloon Analogue Risk Task, BDI = Beck Depression Inventory, MINI = Mini-International Neuropsychiatric Interview, SCID = Structured Clinical interview, CESD= Center for Epidemiological Studies Depression Scale, ICD = International Classification of Diseases, version 10 (ICD-10).

| Author | Disorder | Instrument | Task | Outcome Measure |
| --- | --- | --- | --- | --- |
| Gao et al., 2021 | MDD | SCID-I (DSM-IV-TR) | BART task | Average number of pumps |
| Bi et al., 2022 | Subthreshold depression | Beck Depression Inventory–II (self-report) | Effort expenditure task | Prosocial effort-based decision-making: proportion of effortful choices for others |
| Smoski et al., 2008 | High vs. low depressed traits(cut-off group assignment ) | Hamilton Depression Rating Scale (self-report) | IGT | Selection of risky desks |
| Jollant et al., 2016 | MDD | SCID-IV (DSM-IV-TR) + HAMD-21 (cutoff-based) | IGT | net score: advantageous minus disadvantageous |
| Gu et al., 2020 | MDD | MINI | IGT | Net scores |
| Kong et al., 2022 | MDD (first-episode) | SCID-IV | BART | mean pumps on win balloons |
| Ji et al., 2021 | MDD | SCID-IV | Bart task | mean pumps on win balloons |
| Saperia et al., 2019 | MDD | SCID-IV | IGT | advantageous deck selections |
| Ortega et al., 2017 | High vs. low depressive traits (STAI-T + BDI; cutoff-based) | BDI and STAI (self-report) | BART | Average number of inflation/pumps |
| Hevey et al., 2017 | MDD | SCID-5 | BART | number of pumps |
| Liu et al., 2022 | MDD | SCID-IV | BART | mean number of pumps of win-balloons |
| Maddox et al., 2012 | High vs. low depressive traits (cut-off group assignment) | CES-D (self-report) | Mars farming task | History-dependent decision-making (gain-maximization): total points |
| Cella et al., 2010 | MDD | SCID-IV | IGT (Phase 1-standard) | Net score |
| Ho et al., 2018 | MDD | MINI | IGT | Net score |
| Kunisato et al., 2012 | High vs. low depressive traits (cut-off group assignment) | CES-D | probabilistic selection task developed by Frank et al. (2004) | Probability of correct selection of for a favorable contingency, Choice A. |
| Mansur et al., 2019 | MDD | MINI | Effort-based decision-making | proportion of hard task choices |
| Zheng et al., 2022 | MDD | SCID-IV | IGT | net total score |
| Subramaniapillai et al., 2019 | MDD | MINI | effort based decision making | proportion of hard task choices |
| Mukherjee et al., 2020* | MDD | SCID-IV | reward learning task | percentage of Rich choices |
| Destoop et al., 2012 | MDD | SCID-IV | Ultimatum Game | Acceptance rates |
| Wang et al., 2014 | MDD | clinical interview based on DSM-IV criteria | Ultimatum Game | acceptance rates of most unfair offers |
| Faulkner et al., 2022 | Depression | MINI | reward-based decision-making task (Huys et al. 2012) | total money won |
| Radke et al., 2013 | MDD | SCID (DSM-IV-TR) | Ultimatum game | Rejection rate of unfair offers: |
| Moniz et al., 2016 | MDD | MINI | IGT | net score |
| Carbajal et al., 2017 | MDD | clinical interview based on DSM-IV criteria | Ultimatum Game | rejection rates |
| Sánchez-Loyo et al., 2016 | MDD | clinical interview based on DSM-IV criteria | Decision-making in social context | Total gain |
| von Helversen et al., 2011 | High vs. low depressive traits (median split assignment) | Patient Health Questionnaire (PHQ-D) | sequential choice task (secretary problem) | Performance: the mean number of points earned per game (i.e., performance) |
| Sande et al., 2019 | MDD | SCID-IV | sequential choice task (secretary problem) | the mean number of points earned per game (i.e., performance) |
| Gorlyn et al., 2013 | MDD | SCID-IV | IGT | net total score |
| Rutledge et al., 2017 | MDD | existing MDD patients from psychological and medical facilities | probabilistic reward task with risk | task earnings |
| Deisenhammer et al., 2018 | MDD | met ICD-10 criteria | IGT | net scores |
| Keskin-Gokcelli et al., 2024 | MDD | SCID-5 | Probabilistic reward learning task | amount earned |
| Zhang et al., 2012 | MDD | MINI | trust game (adapted from McCabe et al. 2001) | deceptive responses (social risk-taking) |
| Fan et al., 2021 | MDD | clinical interview based on DSM-IV criteria | BART | mean pumps on win balloons |
| Lawlor et al., 2020 | MDD | SCID-IV | Probabilistic Reward Task (PRT) | Response bias |
| Ang et al., 2023 | MDD | SCID (DSM-IV-TR) | Cognitive Effort Motivation Task | Proportion of High effort High Reward choices |
| Zou et al., 2020 | MDD | DSM-IV criteria | adapted version of Effort Expenditure for Reward Task. | percentage of high-effort choices. |
| Shao et al, 2015 | MDD | clinical interview based on DSM-IV criteria | modified Trust game | Proportion of cheating choices |
| Must et al., 2006 | MDD | MINI | IGT (standard- ABCD form) | net scores, block 5 (last block) |
| Mukherjee et al., 2020** | MDD | SCID-IV | probabilistic reversal-learning task | proportion rich choices, reward condition |
| Benke et al., 2021 | MDD | ICD-10 diagnosis of major depression | Probability-Associated Gambling Task (PAG) | total winning amount |
| Zhang et al., 2023 | MDD | SCID-IV | BART | Risk-taking Behavior score |

**Supplementary Table 4.**
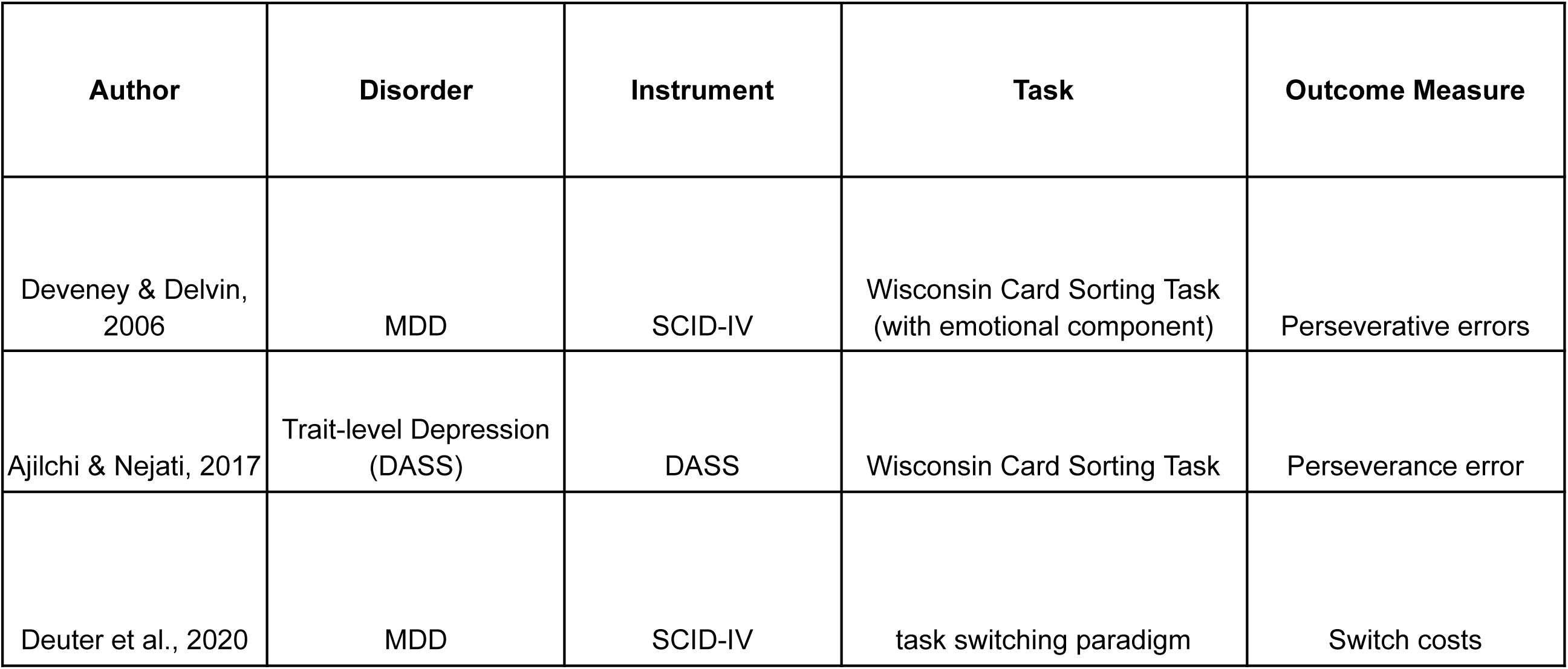
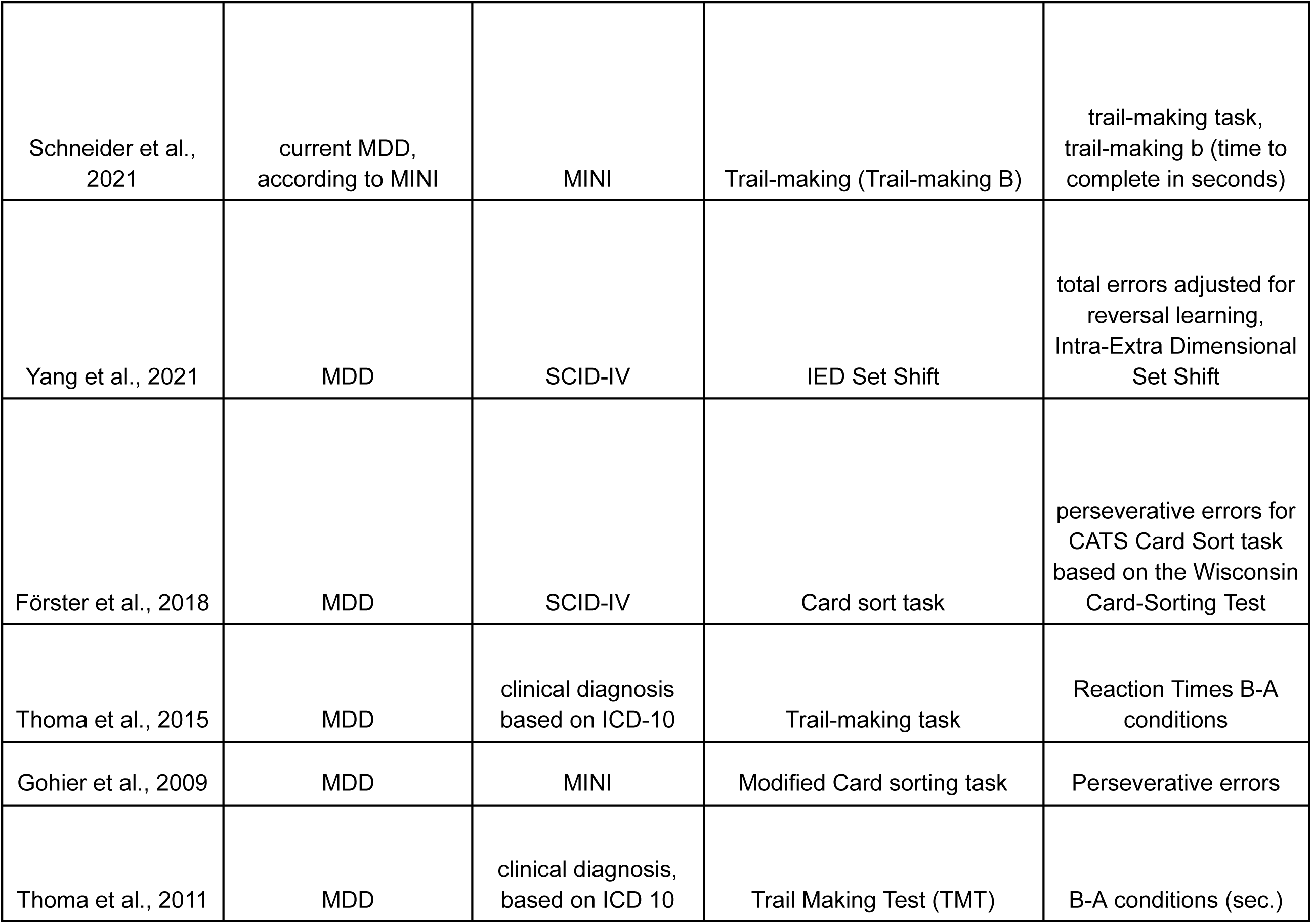
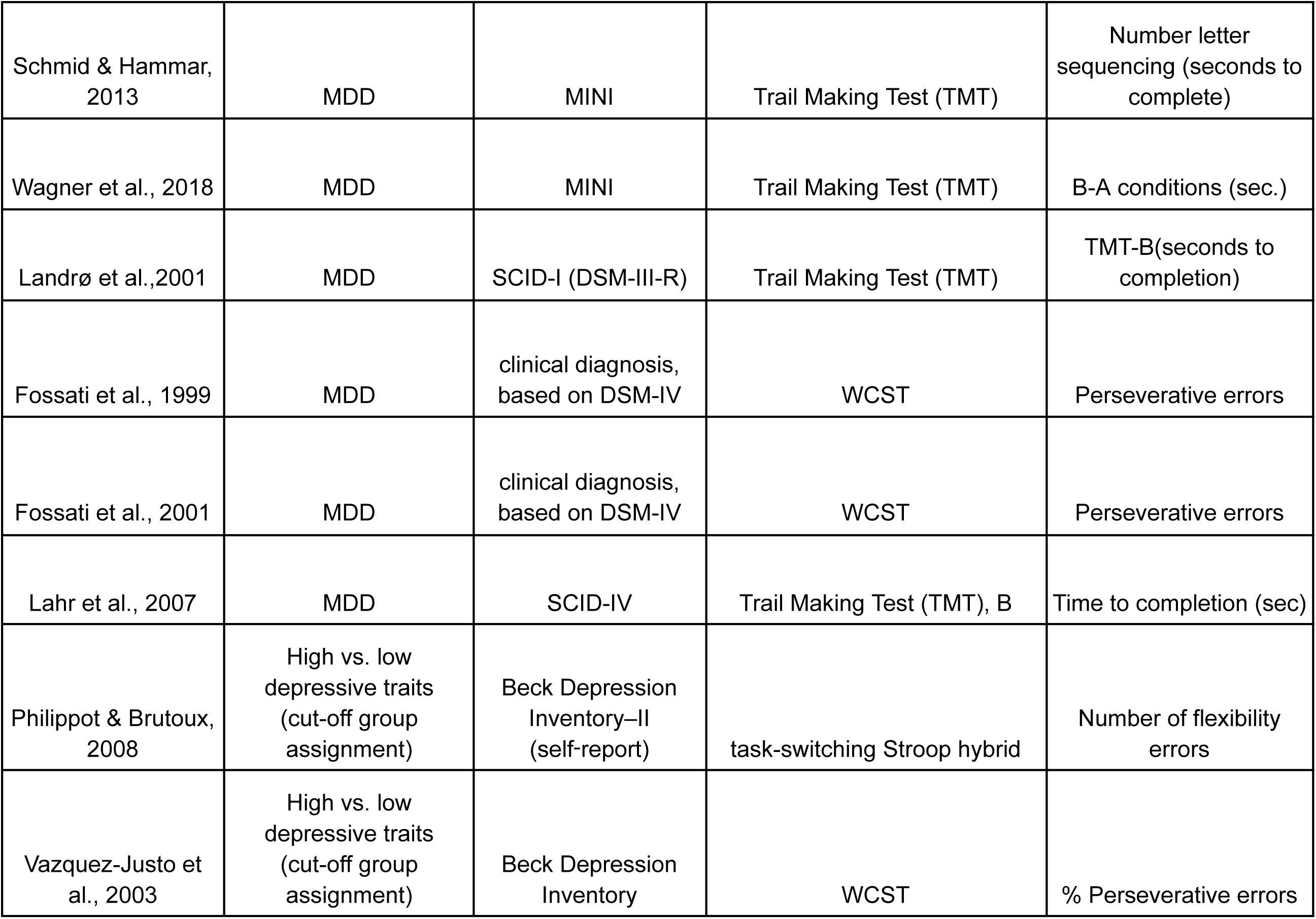
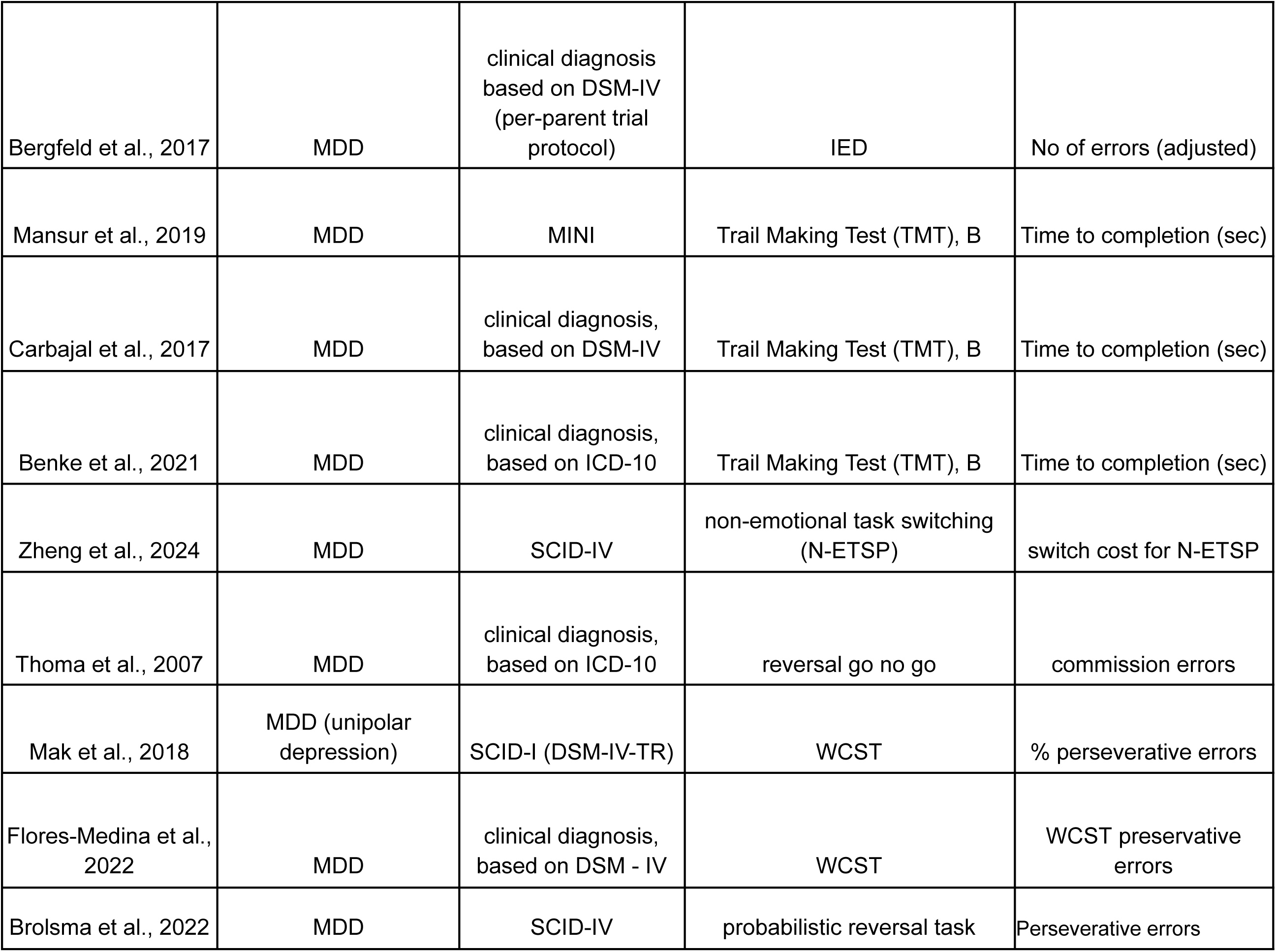
Effects of Depression on flexibility: Summary of the characteristics of articles included in the meta-analysis for the full model ( includes articles that were established as outliers).Abbreviations MINI = Mini-International Neuropsychiatric Interview, SCID = Structured Clinical interview, MDD= major depressive disorder.

**Supplementary Table 5:**
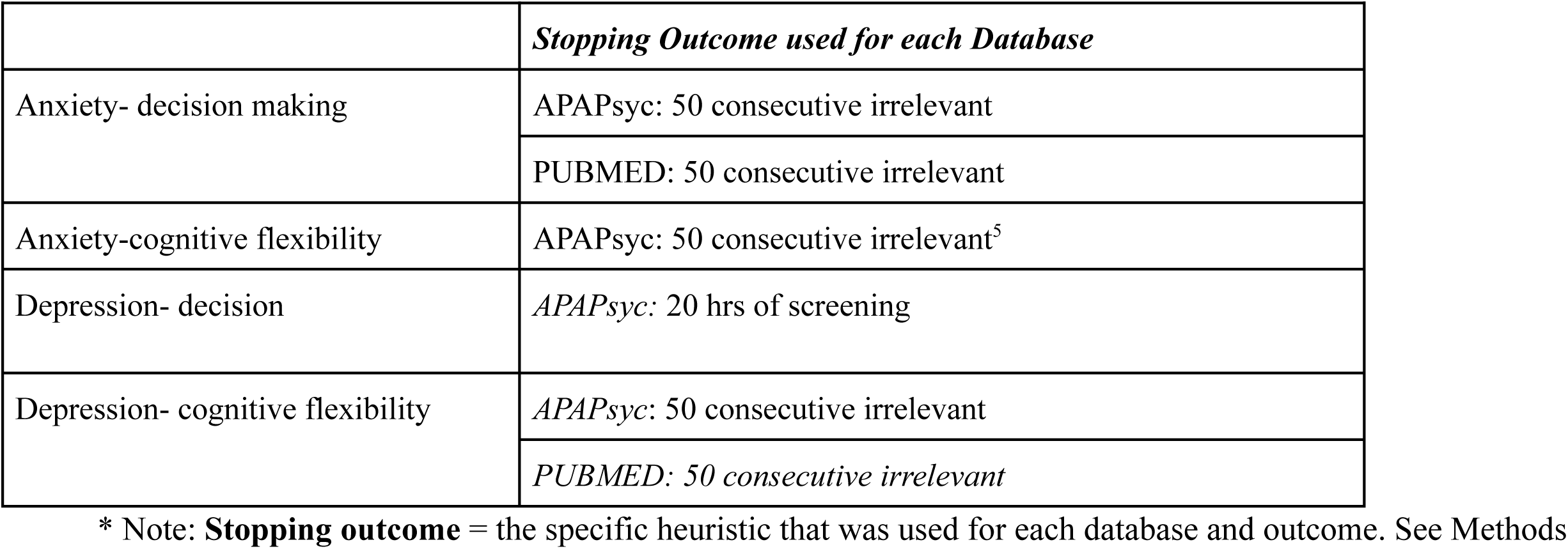
AI-assisted Abstract Screening.

## Supplementary Figures

**Supp Figure 1A:**
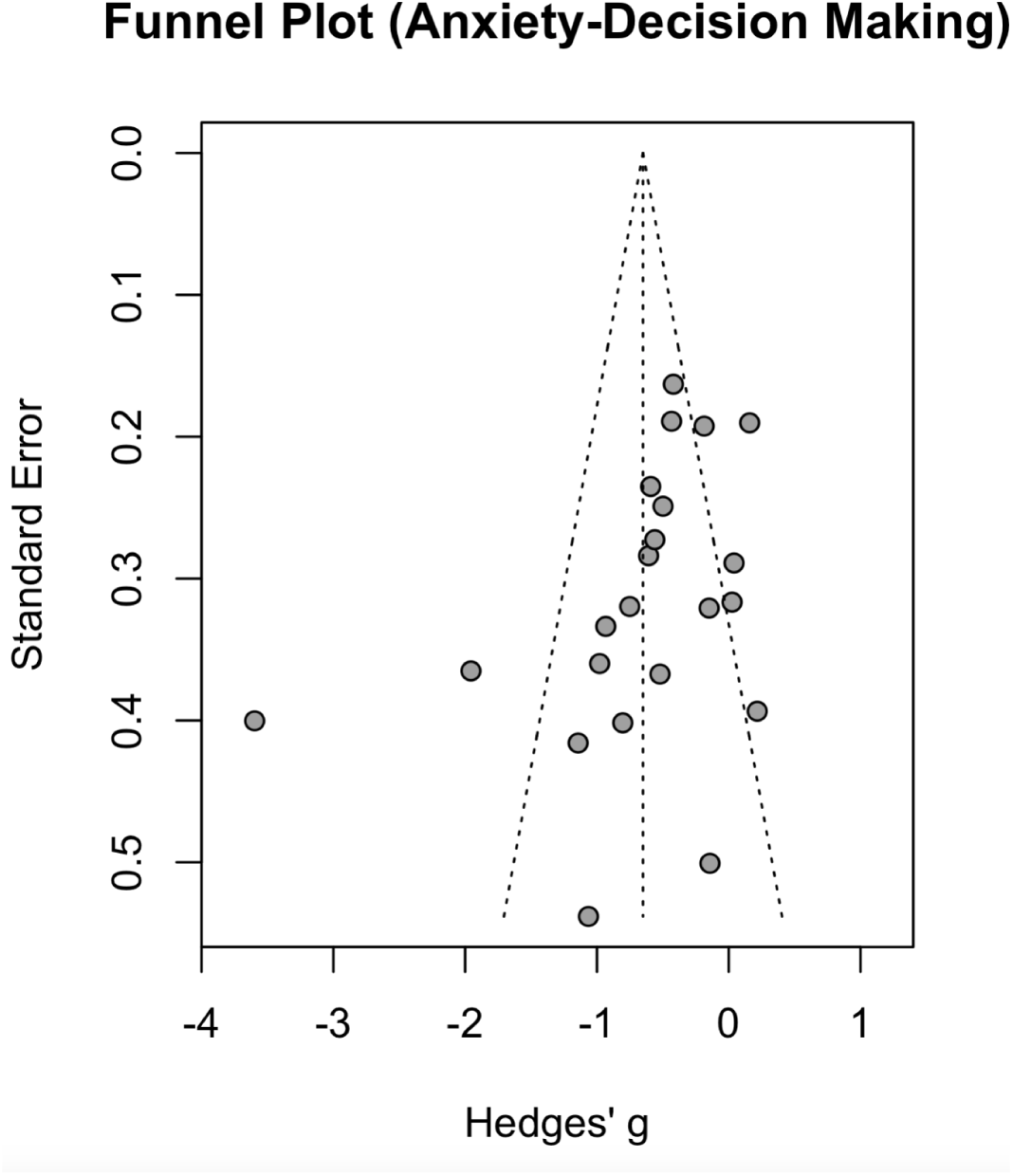
Funnel plot for the full meta-analytic model examining anxiety and decision-making. Each point represents an individual study’s Hedges’ g plotted against its standard error. The vertical line marks the pooled effect estimate, and the dashed lines indicate the expected 95% confidence region of the funnel.

**Supp Figure 1B:**
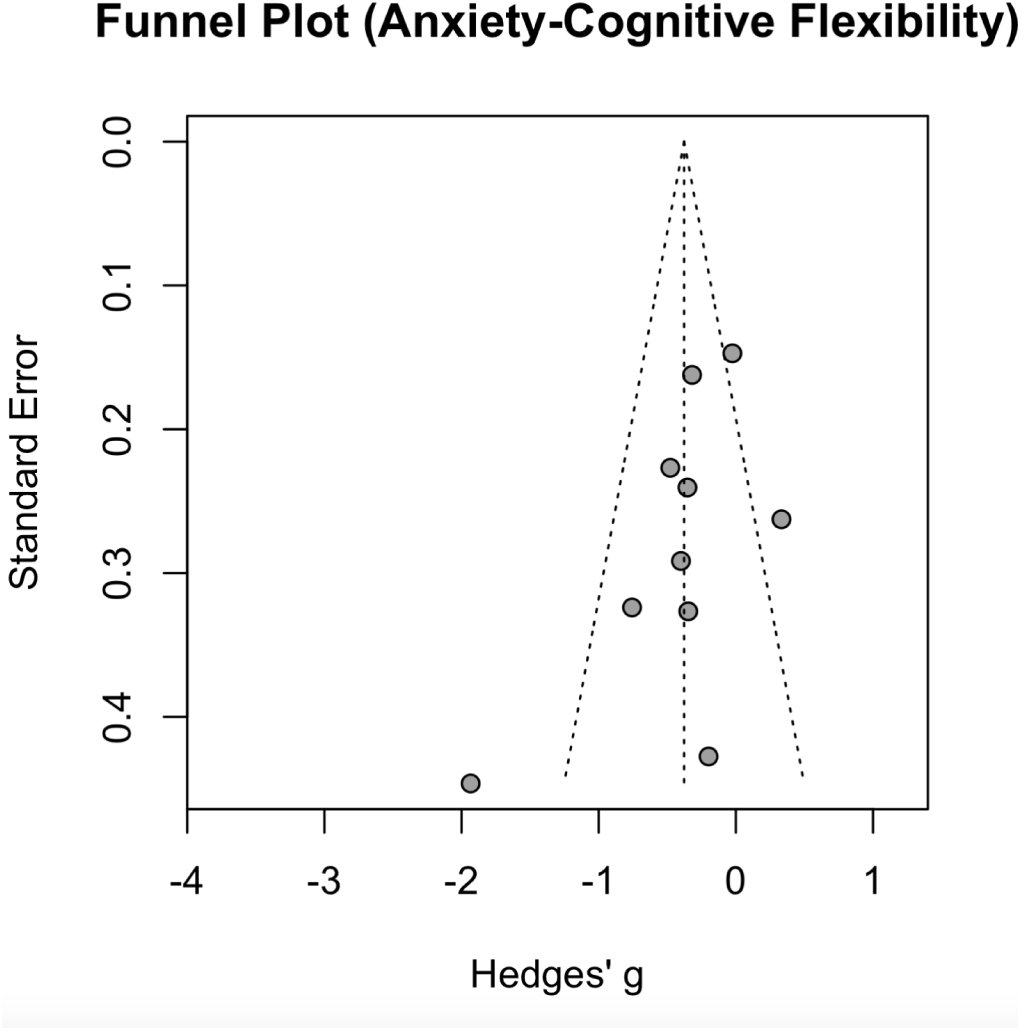
Funnel plot for the full meta-analytic model examining anxiety and cognitive flexibility. Each point represents an individual study’s Hedges’ g plotted against its standard error. The vertical line marks the pooled effect estimate, and the dashed lines indicate the expected 95% confidence region of the funnel.

**Supp Figure 1C:**
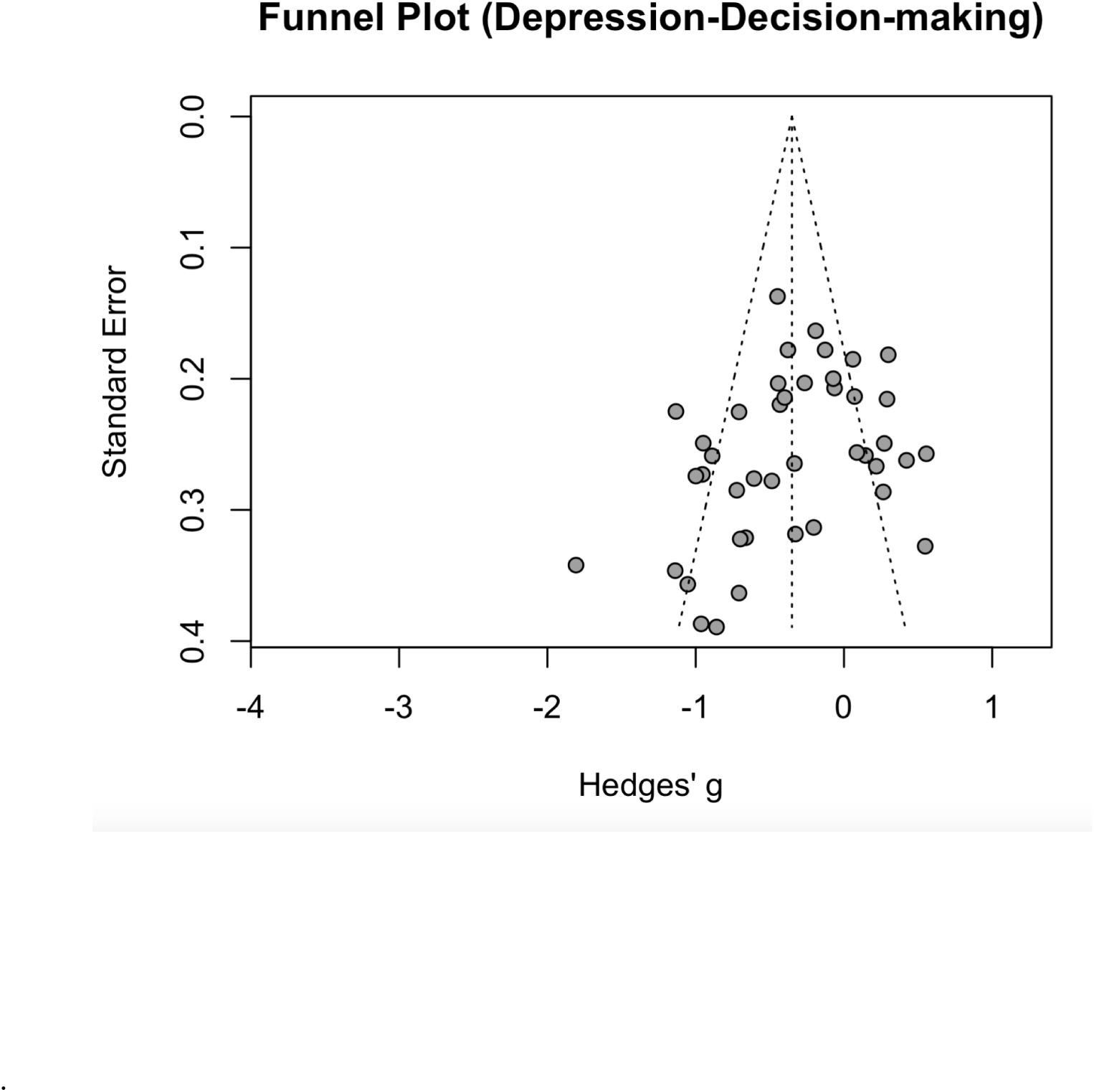
Funnel plot for the full meta-analytic model examining depression and decision-making Each point represents an individual study’s Hedges’ g plotted against its standard error. The vertical line marks the pooled effect estimate, and the dashed lines indicate the expected 95% confidence region of the funnel.

**Supp Figure 1D:**
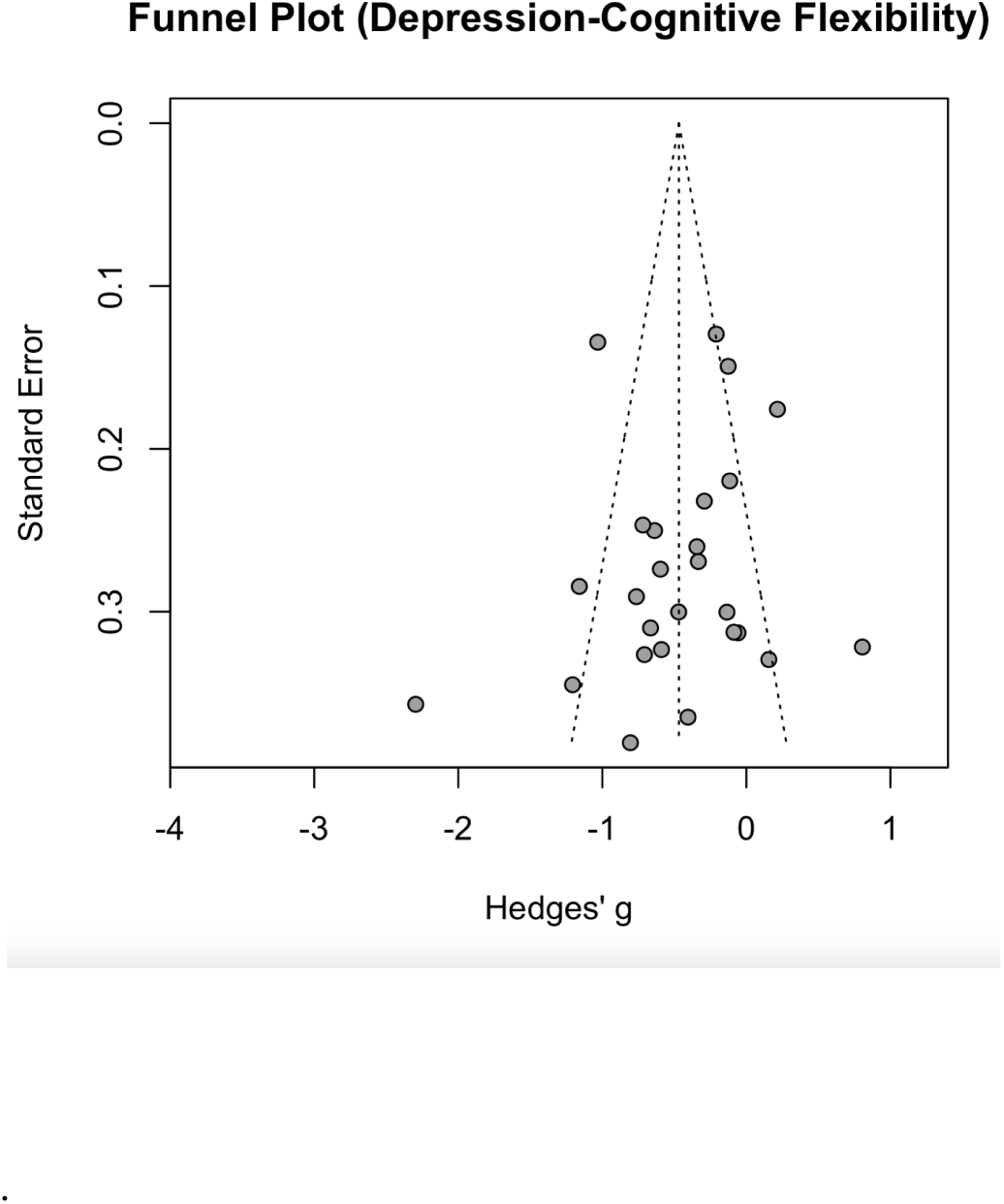
Funnel plot for the full meta-analytic model examining depression and cognitive flexibility. Each point represents an individual study’s Hedges’ g plotted against its standard error. The vertical line marks the pooled effect estimate, and the dashed lines indicate the expected 95% confidence region of the funnel.

## Appendix

***List of studies included for meta-analysis (outliers included)***

## Footnotes

1 Studies reporting clinical diagnoses of anxiety and depressive disorders were included if the study investigated a disorder defined by the DSM V as an anxiety disorder or depressive disorder.

2 While other meta-analyses may use custom publication date ranges for queries, we allowed for an exhaustive search by not using custom publication date ranges.

3 ASReview documentation stipulates a minimum training data set of 2, with at least one relevant and one irrelevant labeled record.

4 ASReview documentation stipulates a minimum training data set of 2, with at least one relevant and one irrelevant labeled record.

5 PUBMED database for anxiety-cognitive flexibility was completed without AI-assisted screening.

